# An interpretable and interactive clinical AI agent for personalized anti-infective decision support in carbapenem-resistant Gram-negative bacterial infection

**DOI:** 10.64898/2026.05.18.26353005

**Authors:** Xiwen Cao, Dake Shi, Zhaohui Du, Jiaxuan Zhou, Zhe Wang, Zhe Liu, Qi Wang

## Abstract

Carbapenem-resistant Gram-negative bacteria (CRGNB) infections remain difficult to manage because treatment decisions must balance heterogeneous patient risk, limited antibiotic options, potential toxicity and emerging resistance. Clinical care in this setting requires not only single-endpoint risk prediction, but also decision-support frameworks that can jointly enable prognosis assessment, result interpretation, and individualized treatment comparison. Here we present Dr.BUG, an interactive clinical AI agent for personalized decision support in CRGNB infection. Dr.BUG integrates stable feature-set selection, multi-task prognostic modelling, interpretability analysis and model-based simulation of antibiotic regimen recommendation into a unified workflow. Using a development cohort, a temporally independent validation cohort, and external cohorts from the MIMIC-IV dataset, we developed and validated models for four clinically relevant tasks: clinical efficacy, survival outcome, polymyxin resistance and treatment duration. Model inputs were derived primarily from routinely available and relatively low-cost clinical variables, supporting translational feasibility. Across the major tasks, selected-feature models matched or exceeded the performance of their full-feature counterparts while using fewer variables, as reflected in 82.0% of optimized-metric comparisons in the development cohort, and remained robust in both temporal and external validation. Dr.BUG further provided both population-level and patient-level interpretability and generated individualized rankings of candidate antibiotic regimens. In the retrospective analysis of non-survivors, clinician review suggested that regimens recommended by Dr.BUG might be associated with higher predicted survival probabilities. These findings support a broader role for clinical AI in complex drug-resistant infections, extending its utility from offline risk prediction to interpretable, deployable, and personalized decision support.

## Introduction

Antimicrobial resistance has become a major global public health threat[1]. It continues to compromise the treatability of bacterial infections and increases the risks of death, prolonged hospitalization and rising healthcare costs[2–8]. In hospital-acquired and critical-care settings, carbapenem-resistant Gram-negative bacteria (CRGNB) have emerged as a major challenge for anti-infective management because of limited treatment options, substantial patient heterogeneity and poor outcomes[9–11]. Common high-risk pathogens include carbapenem-resistant *Enterobacteriaceae* (CRE), carbapenem-resistant *Acinetobacter baumannii* (CRAB) and carbapenem-resistant *Pseudomonas aeruginosa* (CRPA)[11, 12]. Among these, CRAB remains classified as a critical priority pathogen in the 2024 updated World Health Organization bacterial priority pathogens list[12, 13].

CRAB infections are common among hospitalized and critically ill patients and can cause severe healthcare-associated infections, including pneumonia, bloodstream infection and urinary tract infection[14–17]. CRAB is particularly difficult to treat, partly because recent antimicrobial development has offered comparatively few effective options for this pathogen[11, 16]. Clinical outcomes are shaped not only by pathogen resistance, but also by host status, infection severity, organ function and actual treatment exposure[11, 16, 18, 19]. Clinicians still rely on polymyxins-containing regimens[16, 20]. However, the clinical use of polymyxins is constrained by their narrow therapeutic window, complex pharmacokinetic and pharmacodynamic properties, and limited routine monitoring in clinical practice, making dose optimization difficult in routine care[20–22]. CRAB therefore represents a stringent test case for individualized anti-infective decision support, with potential methodological relevance to other CRGNB infections, such as CRE and CRPA[11, 16, 20].

According to the core principles of antimicrobial stewardship (AMS)[23], the treatment of complex drug-resistant infections requires not only appropriate antibiotic selection, but also regimen optimization to improve patient outcomes while minimizing toxicity and further selection pressure for resistance[24, 25]. This need is particularly clear in severe CRAB infection, where patient status may change rapidly and early treatment decisions are often made before complete microbiological and laboratory information is available[11, 16, 26]. Clinicians therefore frequently make empirical decisions under uncertainty and must revise treatment as new information becomes available[23, 26, 27]. These features expose the limitations of decision support tools that provide only static rules or isolated risk estimates, and support the need for systems that integrate patient status, treatment exposure and model-derived explanations to enable iterative, individualized comparison of candidate treatment strategies[27–31].

Machine learning offers new opportunities for prediction and decision support in drug-resistant infections[32–34]. Previous studies have used electronic health records and clinical variables to predict the appropriateness of empirical antibiotic therapy, the risk of adverse outcomes and the probability of resistance[35–39]. Some have also attempted to compare the potential utility of different antibiotics in specific infectious scenarios[31, 39, 40]. However, from a translational perspective, several key limitations remain[30, 33]. First, feature selection in clinical prediction studies is commonly tied to a specific modelling strategy[41–44]. As a result, the derived feature sets may lack sufficient stability, cross-model consistency and transferability across heterogeneous datasets[41, 42, 44]. Common approaches include statistical prescreening, recursive feature elimination, and variable selection based on LASSO, tree-based feature importance or SHAP ranking[42, 43, 45]. Second, many existing models still rely mainly on static patient characteristics and do not adequately account for treatment intensity, dosing regimens or actual medication differences[32–34, 46]. This limits their ability to support more granular and individualized anti-infective decision-making[33, 34, 46]. Third, most studies remain at the stage of model development or offline evaluation. They have not yet been translated into low-barrier tools that clinicians can readily use, invoke and extend in real-world practice[27, 30, 33, 34].

Against this background, agentic AI and interactive clinical AI may provide a practical path towards closing this translational gap[47–49]. Their central value lies in whether validated modules for prediction, interpretation and treatment support can be organized around specific clinical goals into coherent, traceable and controllable workflows[27, 29, 47, 48]. In infection management, clinical needs usually extend beyond a single risk score[27, 31, 33, 34]. They also include effective links between outcome assessment, result interpretation and comparison of candidate treatment strategies[27, 29, 31, 34]. Therefore, the key issue is not whether a system takes the form of an “agent”, but whether these modules can be integrated into a clinically usable and deployable framework[27, 30, 47, 48].

In this study, we developed Dr.BUG, an interactive clinical AI agent for personalized anti-infective decision support in CRGNB infections, using CRAB as the principal representative pathogen. To address the strong redundancy and limited stability of variables in high-dimensional clinical data, we proposed a feature-set selection strategy that emphasizes cross-model stability and transferability, with the aim of identifying more robust clinical factors. We then incorporated treatment-intensity and medication-related information to build prediction models for clinical improvement, survival outcome, polymyxin resistance and treatment duration. These models were systematically evaluated in a development cohort, a temporal validation cohort and two external cohorts from the MIMIC-IV dataset[50]. We further integrated patient-level prediction, result interpretation and individualized treatment recommendation into a unified interactive system to support more interpretable and traceable individualized anti-infective decision-making. By allowing clinicians to train models on datasets defined by specific clinical time windows, Dr.BUG provides a practical route towards dynamic, stage-specific prediction as disease status and treatment exposure evolve. Overall, this study presents a clinical AI framework for the individualized management of complex drug-resistant infections and provides a methodological basis for building interpretable, deployable and clinically translatable decision-support tools in the CRGNB setting. It also provides a practical example of how agentic and interactive AI can be adapted to infectious-disease care, and may inform the development of similar decision-support frameworks for other complex clinical scenarios requiring individualized treatment decisions.

## Results

### Study overview

CRGNB infections remain difficult to manage in routine care because patient outcomes and treatment responses are highly heterogeneous. There is therefore a need for practical digital tools to support individualized anti-infective decision-making. In this study, Dr.BUG was developed as an interactive clinical AI agent for personalized anti-infective management of CRGNB infections, integrating model development, patient-level prediction, interpretability analysis and individualized antibiotic recommendation within a unified workflow (Figure 1). The framework was designed to support a broader clinical decision process, including model construction and updating, personalized outcome assessment, explanation of model outputs and reference for treatment selection.

**Figure 1.**
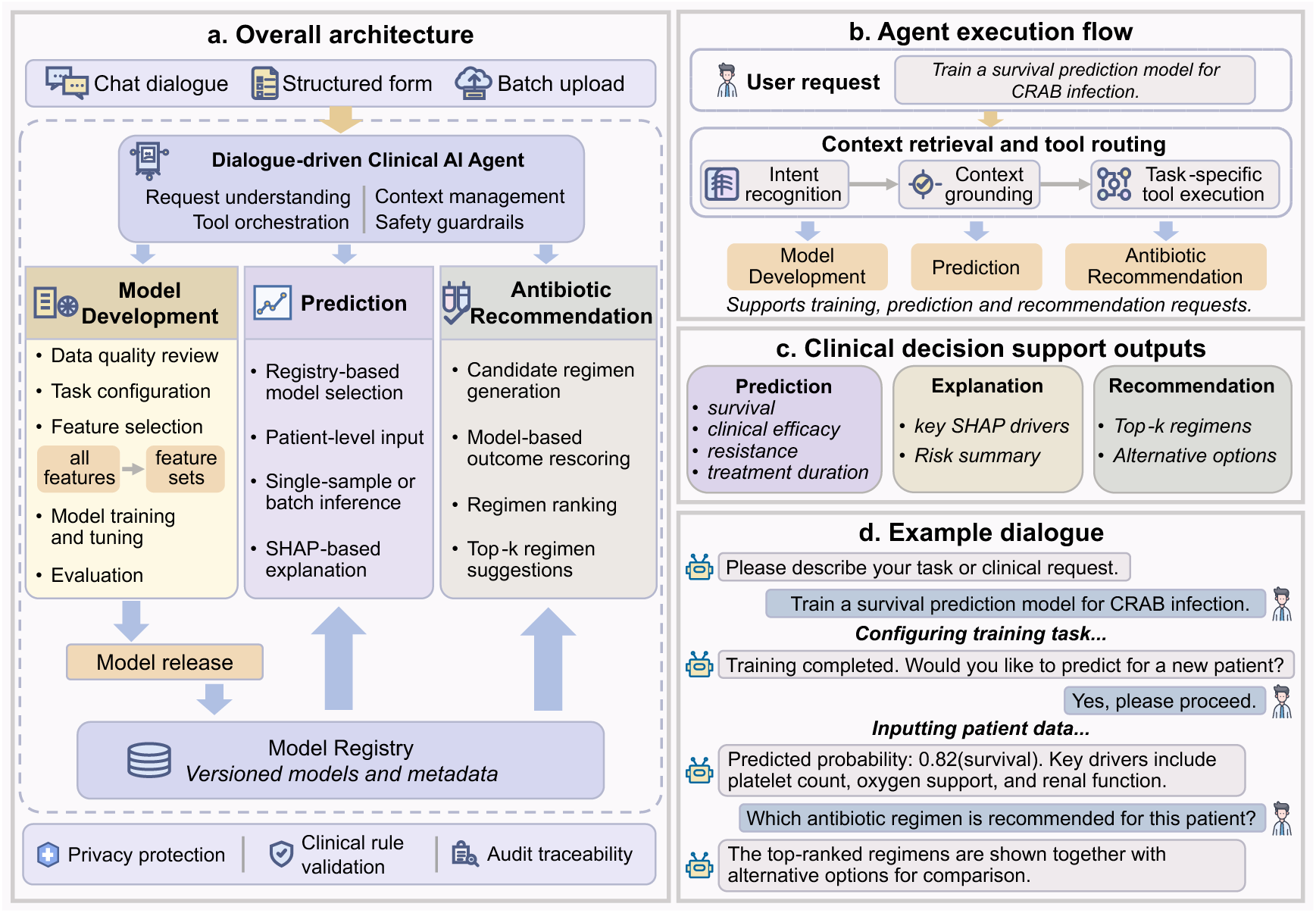
Framework of Dr.BUG. (a) Overall architecture of Dr.BUG. The system uses dialogue as a unified entry point and supports model development, patient-level prediction and antibiotic recommendation; trained models are released into a versioned model registry for downstream use. (b) Agent execution flow. User requests are processed through intent recognition, context grounding and tool invocation, enabling training, prediction or recommendation within a unified workflow. (c) Clinical decision-support outputs. Dr.BUG provides predictions for survival, clinical efficacy, resistance and treatment duration, together with SHAP-based explanation and ranked candidate antibiotic regimens. (d) Example dialogue. An end-to-end interaction is shown, spanning task initiation, model training, patient-level prediction and regimen recommendation.

We organized the subsequent analyses as a hierarchical evidence chain for the core functions of Dr.BUG (Figure 2). We focused on four clinically relevant tasks in anti-infective management, including prediction of clinical efficacy, survival outcome, polymyxin resistance and treatment duration (Figure 2b). In the development cohort, we first implemented a multi-model voting-based framework to identify feature subsets that preserved both predictive performance and stability (Figure 2c). We then combined these analyses with SHAP[45] profiling to assess whether the resulting models remained clinically interpretable. Temporal validation was performed to examine robustness under evolving clinical conditions. A two-level multicenter evaluation was performed using the MIMIC-IV dataset (Figure 2d). In the external CRAB cohort, we assessed the cross-center generalizability of the selected feature sets. In the independent CRPA cohort, we further examined whether the feature-selection strategy itself could be transferred to another carbapenem-resistant pathogen. Finally, we conducted a simulation study of individualized antibiotic recommendation, extending the framework from risk estimation to treatment comparison (Figure 2e). These validated components were ultimately integrated into Dr.BUG.

**Figure 2.**
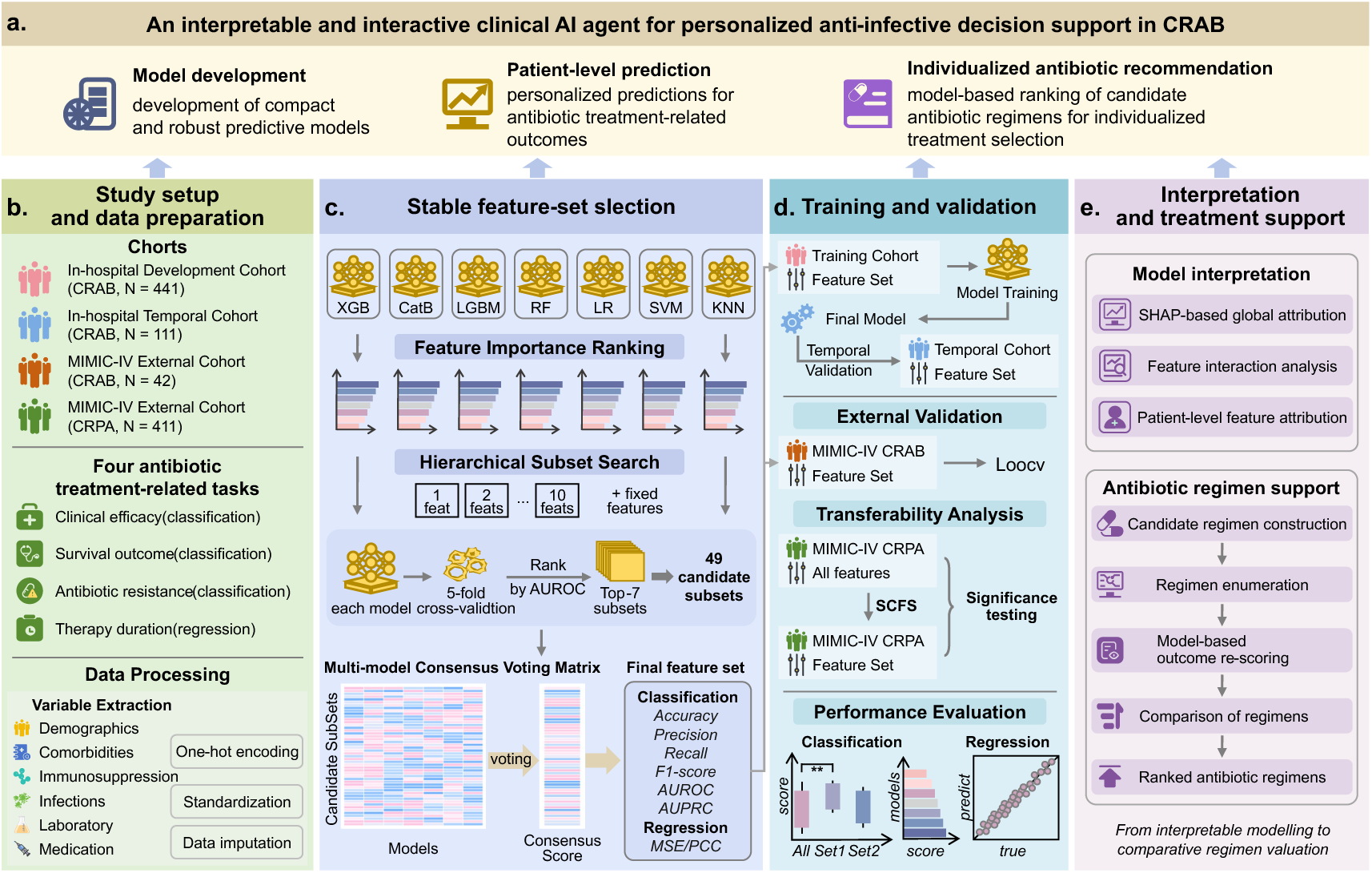
Overview of the study workflow. (a) Overall framework of Dr.BUG. (b) Study setup and data preparation, including cohort construction, definition of four antibiotic treatment-related prediction tasks, variable extraction, and data preprocessing. (c) Stable feature-set selection workflow. In the development cohort, features were ranked according to model-specific mean absolute SHAP values, and the top 10 features were retained as the candidate pool. Stable feature subsets were then identified through hierarchical subset search and multi-model consensus voting. (d) Model training and layered validation across the development cohort, temporal validation cohort, and external cohorts from the MIMIC-IV dataset, together with performance evaluation for both classification and regression tasks. (e) Model interpretation and antibiotic treatment support, including interpretability analysis based on SHAP and antibiotic recommendation simulation.

### Cohort characteristics

The in-hospital evaluation was based on a development cohort and a temporally independent validation cohort from the same center. The development cohort comprised 441 hospitalized patients from Ruijin Hospital, Shanghai Jiao Tong University School of Medicine, who were diagnosed with CRAB infection and received systemic antibiotic therapy between October 2020 and September 2024. An additional 111 incident cases from the same center, enrolled between October 2024 and May 2025, were used to construct the temporal validation cohort. The development and temporal validation cohorts were broadly comparable in baseline characteristics, including demographics, overall clinical status and comorbidity burden, with no statistically significant differences observed (all *P* > 0.05; Supplementary Table 1). In the development cohort, 327 patients achieved clinical improvement and 114 experienced treatment failure; the corresponding numbers in the temporal validation cohort were 90 and 21. Both cohorts were predominantly composed of ICU patients, and most individuals received polymyxin-containing combination regimens (Supplementary Table 1).

In total, we extracted 22 continuous variables and 25 categorical variables. The continuous variables included measurements available before CRAB diagnosis and treatment initiation, together with the worst values from 2 days before to 2 days after CRPA positivity. In addition, 10 antibiotic-related features were incorporated for subsequent modelling.

### Feature subsets improve predictive performance and clinical translatability

Clinical decision support is more likely to be deployable when predictive performance can be maintained with a compact and clinically tractable input space. We therefore compared the performance of the model trained on the entire feature set (Full-Feature Model, FFM) with the model utilizing the subset identified by our feature-set selection strategy (Selected-Feature Model, SFM) across four antibiotic treatment-related tasks in the development cohort. Within each task, feature sets were selected according to the optimization metrics. Each SFM was compared with its corresponding FFM using the same metric that guided feature-set selection. Across four tasks, SFMs matched or exceeded FFMs in 109 of 133 comparisons based on these optimized metrics (82.0%). Performance differences between FFMs and SFMs were further assessed within classification tasks using paired statistical testing across model-fold evaluations. The optimized feature subsets and their constituent variables for each task are summarized in Supplementary Table 2. Antibiotic-treatment variables were retained as mandatory inputs for survival prediction, clinical efficacy prediction and treatment-duration prediction.

The clearest benefit of the SFMs was observed in survival prediction. Two representative subsets were identified: Survival feature-set A, optimized for Precision and AUROC, and Survival feature-set B, optimized for Accuracy, Recall, F1-score and AUPRC. Compared with FFMs, these SFMs showed a consistent upward shift across all classification metrics. The strongest gains were seen in the metrics used to define each subset. The subset optimized for Accuracy, Recall, F1-score and AUPRC significantly improved all four measures (Figure 3a; Accuracy, P<0.001; Recall, P<0.05; F1-score, P<0.001; AUPRC, P<0.01). The subset optimized for Precision and AUROC produced even stronger improvements in both target metrics, with P<0.001 for each. Notably, this benefit was observed even though the FFMs already performed well. CatBoost-FFM and RF-FFM both reached AUROC values of approximately 0.76, and AUPRC approached 0.89. Performance improved further after restriction to the optimized subset. Using the subset optimized for Precision and AUROC as an example, CatBoost-SFM achieved an AUROC of 0.791 and an AUPRC of 0.905, SVM-SFM achieved an F1-score of 0.865 and a Recall of 0.954, and RF-SFM achieved a Recall of 0.951 (Figure 3c-d). Aside from minor fluctuations in isolated metrics for a small number of models, the vast majority of performance indicators improved in a concordant manner. These results indicate that the prognostic signal required for survival prediction does not depend on a high-dimensional input space, but can be captured more effectively by a smaller set of more informative clinical variables. The detailed performance metrics of each model across the different feature subsets are shown in Supplementary Figure 1.

**Figure 3.**
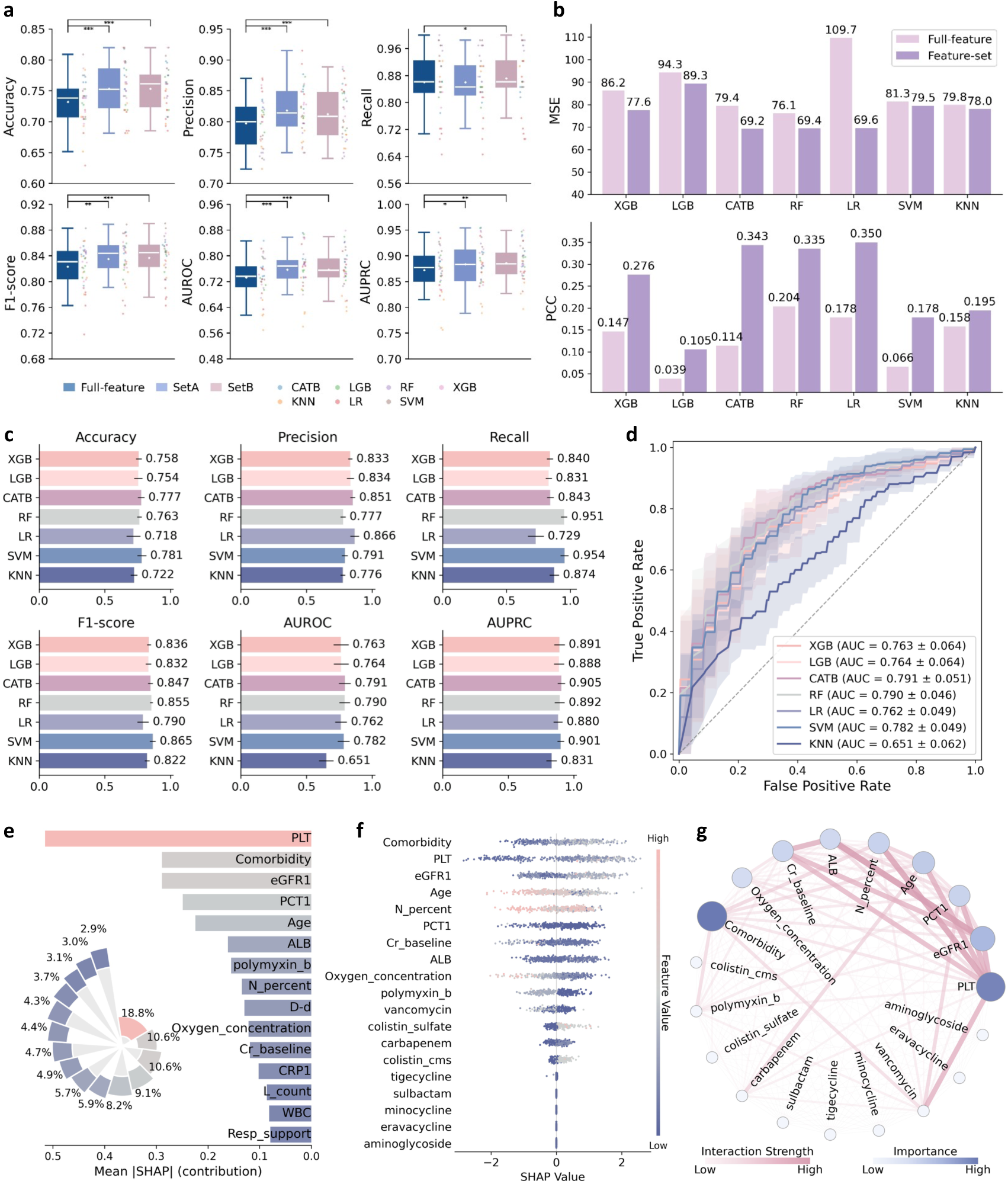
Model performance in the development cohort and interpretability analysis of a representative survival model. (a) Fivefold cross-validation comparison between FFMs and SFMs for survival prediction. Dots in the boxplots indicate results from individual models across cross-validation folds; boxes represent the interquartile range and center lines denote the median. Statistical significance was assessed using one-sided paired Wilcoxon signed-rank tests: ∗ *P* < 0.05, ∗∗ *P* < 0.01, and ∗∗∗ *P* < 0.001. (XGB Extreme Gradient Boosting, LGB Light Gradient Boosting Machine, CATB Categorical Boosting, RF random forest, LR logistic regression, SVM support vector machine, KNN k-nearest neighbors) (b) Performance comparison between FFMs and SFMs for treatment-duration prediction. (c-d) Overall performance of a representative optimized feature subset across different models for survival prediction and the corresponding ROC curves; shaded areas denote standard deviation. (e) Global feature importance ranking of the CatBoost-FFM for survival prediction, showing the top 15 features ranked by mean absolute SHAP value, and the top 10 features were retained in the candidate pool for subsequent feature-set selection. (f) SHAP beeswarm plot of the XGBoost-SFM showing the direction and magnitude of feature contributions to model output. Each dot represents one patient; color indicates feature value from low (blue) to high (red), and the sign of the SHAP value indicates positive or negative contribution to the model output. (g) Feature interaction network of the same XGBoost-SFM. Node size and color intensity indicate global feature importance, and edge width reflects interaction strength between features.

A similar pattern was observed for treatment-duration prediction. Analyses were restricted on clinical grounds by excluding patients who died, were discharged because of critical illness, or experienced clinical failure, resulting in a final analytic cohort of 277 patients. For this task, feature-set selection was optimized primarily using MSE as the target metric. After feature selection, PCC increased and MSE decreased in each model (Figure 3b). The largest improvement was seen in linear regression. LR-FFM yielded an MSE of 109.7 and a PCC of 0.178, whereas LR-SFM reduced the MSE to 69.6 and increased the PCC to 0.350. Similar trends were observed in other models. For CatBoost, MSE decreased from 79.4 to 69.2, with PCC increasing from 0.114 to 0.343. Together, these findings indicate that, even for continuous-outcome prediction, optimized feature subsets can attenuate the adverse effects of noisy variables on model fitting and improve concordance between predicted and observed treatment duration. Similar findings were also observed for clinical efficacy prediction and polymyxin resistance prediction, as detailed in Supplementary Note 1, Supplementary Figure 2, Supplementary Figure 3, Supplementary Note 2, Supplementary Figure 4 and Supplementary Figure 5.

Overall, the advantage of the SFMs was not confined to a single algorithm or a single evaluation metric, but emerged as a broader cross-task and cross-model pattern. In clinically relevant prediction tasks, SFMs consistently maintained or improved performance while reducing input complexity. This benefit was reflected either in statistically significant gains in key metrics, as observed in survival prediction, or in tighter performance distributions and greater robustness across folds.

### Core clinical factors underpin survival risk stratification

We next evaluated which clinical signals were driving survival prediction by examining the global distribution of feature contributions in the FFMs for each task, and characterizing effect directionality and feature interactions within the SFMs (Figure 3e-g).

For survival prediction, the top 15 features ranked by mean absolute SHAP values in the CatBoost-FFM primarily represented comorbidity burden, infection-phase hematological and inflammatory markers, as well as pre-treatment renal function and nutritional status (Figure 3e). The representative feature subsets retained most of these high-contribution variables, indicating that dimensionality reduction preserved the core prognostic information used for survival stratification. In the XGBoost-SFM, older age, higher comorbidity burden, thrombocytopenia, renal impairment, and greater oxygen requirement were all associated with a lower predicted probability of survival (Figure 3f). Clinically, these features point to a common pattern of poorer baseline condition, greater acute illness severity and more advanced organ dysfunction. The interaction analysis further showed that these predictors were tightly connected, indicating that the model did not rely on isolated variables but on a clinically coherent pattern of patient vulnerability and acute illness severity (Figure 3g). All these results suggest that the model captured a clinically coherent vulnerability profile instead of relying on single-variable association. Analogous analyses for clinical efficacy and polymyxin resistance prediction are presented in the Supplementary Note 3, Supplementary Note 4 and Supplementary Figure 6.

These SHAP-based analyses indicate that the SFM preserved the principal prognostic signals identified by the FFM, while maintaining effect directions and interaction structures that were clinically interpretable. This supports the view that the reduced model achieved parsimony without losing clinical coherence. These findings suggest that the SFMs maintained predictive performance while providing greater clinical interpretability and decision transparency than the FFMs.

### Temporal validation supports robustness under evolving clinical conditions

To assess whether the selected feature sets remained informative in later real-world patients, we directly evaluated the development-trained FFMs and SFMs in a temporally independent validation cohort of 111 cases collected from October 2024 to May 2025, without further retraining. Despite the temporal shift and the complete separation of this cohort from model development, the SFMs retained clinically meaningful predictive value, supporting their potential robustness in real-world application.

In the survival prediction task, SFMs showed the strongest temporal robustness (Figure 4a). Although performance still differed across classifiers, most SFMs maintained clinically useful discrimination, with an overall pattern similar to that observed in the development cohort. In most models, accuracy remained above 0.73, F1-score above approximately 0.85, AUROC above approximately 0.70, and AUPRC approached 0.90. These findings indicate that the key prognostic signals for survival could still be captured in the temporal cohort by a smaller and more information-dense set of variables, supporting the out-of-time robustness of the corresponding SFMs.

**Figure 4.**
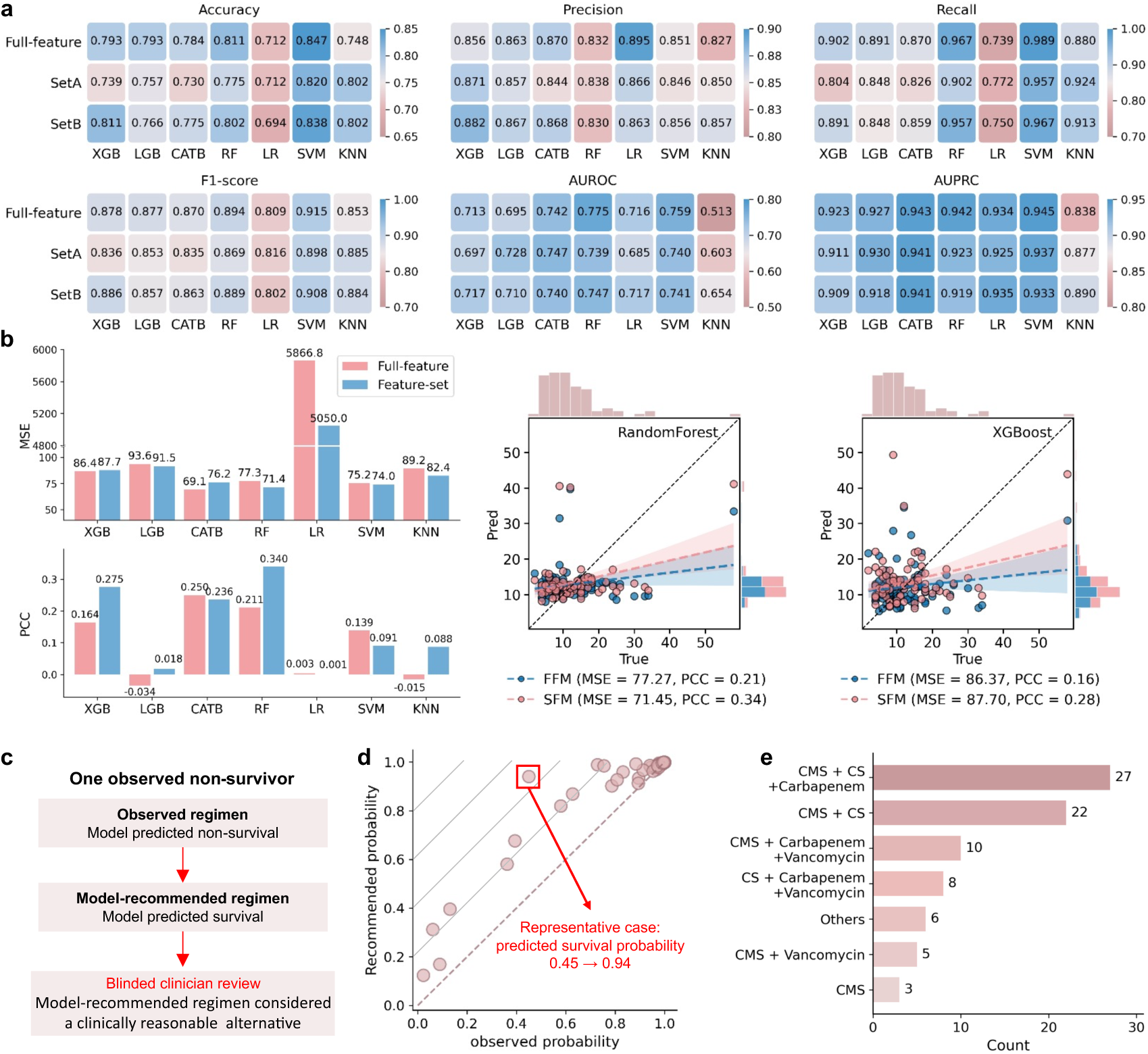
model performance, representative case studies and simulation of individualized antibiotic recommendation in the temporal validation cohort. (a) Survival prediction performance of FFMs and SFMs. (b) Treatment-duration prediction performance across regression models, with representative observed-versus-predicted plots shown for random forest and XGBoost. Marginal histograms show the distributions of observed and predicted values. (c-e) Simulation of individualized antibiotic recommendation for survival prediction. c, Representative observed non-survivor selected for blinded clinical review. The model-recommended regimen was considered clinically reasonable and potentially associated with a higher likelihood of survival. The highlighted point corresponds to the representative case shown in e. d, Predicted survival probabilities under the observed and model-recommended regimens across the 81-patients. e, Frequency distribution of model-recommended regimen backbones.

Treatment-duration prediction showed more limited temporal generalizability (Figure 4b). Compared with FFMs, most SFMs reduced or maintained MSE, but the magnitude of improvement was modest. For example, in random forest, MSE decreased from 77.3 to 71.4 and PCC increased from 0.211 to 0.340. The scatter plot further showed that RF-SFM had a clearer positive trend between observed and predicted treatment duration than RF-FFM, suggesting improved preservation of the relative ordering. However, predictions remained concentrated within a narrow range and underestimated patients with longer observed treatment courses. These findings suggest that the selected feature set can support approximate stratification of treatment duration, but precise estimation of the exact number of treatment days remains difficult in out-of-time validation.

In brief, clinical efficacy prediction retained partial but clinically meaningful discriminative ability in the temporal validation cohort, although its ranking performance was weaker than that in the development cohort. Polymyxin resistance prediction showed greater heterogeneity across models, with feature-set inputs improving resistant-case recognition in some classifiers but not yielding consistent gains overall. Detailed analyses of clinical efficacy prediction and polymyxin resistance prediction are presented in the Supplementary Note 5, Supplementary Note 6 and Supplementary Figure 7. Taken together, these results show that the feature sets identified in the development cohort were able to preserve core predictive signals in prospective real-world patients, particularly for survival prediction, thereby supporting the robustness of the framework under evolving clinical conditions.

### Model-based counterfactual evaluation of regimen recommendation supports personalized treatment evaluation

We next examined whether Dr.BUG could support retrospective treatment re-evaluation. We first focused on a representative non-survivor in the temporal validation cohort. This patient died after receiving the administered antibiotic regimen, suggesting that alternative regimen choices might warrant retrospective consideration. Dr.BUG was then used to enumerate candidate antibiotic regimens and rescore each option using the pretrained survival model. The regimen with the highest predicted probability of survival was defined as the model-recommended regimen.

To assess whether the recommended regimen was clinically reasonable, we invited a clinician to review the patient’s clinical profile together with the administered and model-recommended regimens. The clinician considered the model-recommended regimen clinically reasonable and potentially associated with a higher probability of survival. Consistent with this clinical assessment, the model-recommended regimen increased the model-predicted probability of survival from 45% to 94% (Figure 4c-d). We then extended this counterfactual evaluation to patients in the temporal validation cohort, for whom the pretrained survival model correctly reproduced the observed outcome under the administered regimen, including 7 non-survivors and 74 survivors. Across these patients, the recommended regimen was associated with higher predicted survival probabilities, with varying magnitudes of predicted probability gain across individuals (Figure 4d). Notably, for 3 of the 7 patients who actually died, the model predicted a survival outcome had the recommended regimen been administered. The recommended regimen backbones were distributed across several combinations rather than concentrated in a single fixed pattern (Figure 4e), indicating that Dr.BUG was not constrained to one regimen template and could generate patient-specific recommendations from multiple candidate options.

Taken together, these findings support the potential of the framework as a reference tool for personalized treatment evaluation. By identifying patient-specific alternative regimens associated with higher predicted survival probabilities, the model may help prioritize clinically feasible options for clinician review, particularly in patients with poor predicted prognosis.

### External validation demonstrates cross-center generalizability of both selected feature sets and selection strategy

To assess whether the proposed framework could extend beyond the development center, we performed external validation in two cohorts from the MIMIC-IV dataset (Figure 5). First, we examined whether the survival-associated feature subsets identified in the development cohort could be transferred directly to an independent CRAB population. Second, we examined whether the feature-set selection workflow itself could recover stable predictive subsets in a related but distinct CRPA population. The CRAB cohort established from the MIMIC-IV dataset included 42 CRAB-positive patients who received systemic antibiotic therapy, of whom 33 survived and 9 died. For the survival prediction task, we directly transferred the two feature subsets identified in the development cohort and evaluated them by leave-one-out cross-validation, without repeating feature selection. Despite the small sample size, both transferred subsets retained clinically meaningful discriminative ability. Across models, accuracy was generally around 0.76 to 0.83, and AUPRC ranged from 0.84 to 0.96. LightGBM and logistic regression showed the most stable discrimination. For feature set A, their AUROCs were 0.785 and 0.865, with AUPRCs of 0.917 and 0.958, respectively (Figure 5a). For feature set B, their AUROCs remained 0.721 and 0.828, with AUPRCs of 0.883 and 0.943 (Figure 5b). These two models also identified 7 of the 9 fatal cases under both subsets. By comparison, XGBoost identified 6 of 9 and 5 of 9 deaths under feature sets A and B, and CatBoost identified 3 of 9 deaths under both subsets. Together, these findings indicate that survival-associated feature subsets derived from the development cohort remained informative in an independent external CRAB cohort, supporting their cross-center applicability.

**Figure 5.**
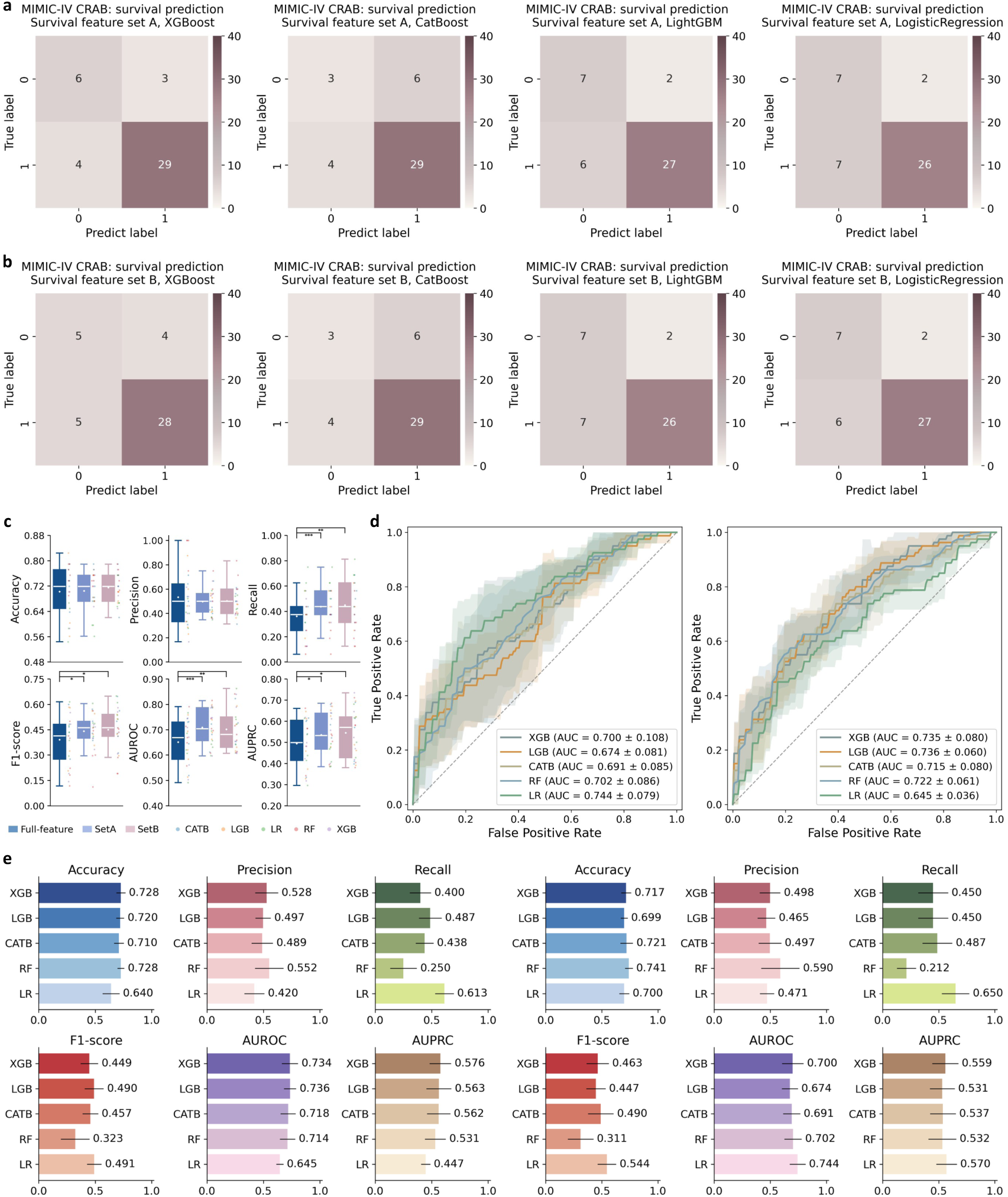
Multicenter validation results in the MIMIC-IV cohorts. (a-b) Two survival-associated feature subsets identified in the development cohort were directly applied to the CRAB cohort from the MIMIC-IV dataset and evaluated using leave-one-out cross-validation, showing the confusion matrices of different models under the two feature subsets. (c) In the CRPA cohort from the MIMIC-IV dataset, the feature-set selection workflow was re-executing using 28-day mortality as the endpoint. Overall performance was compared between FFMs and models built on the two newly selected feature subsets using fivefold cross-validation. (d-e) In the CRPA cohort from the MIMIC-IV dataset, ROC curves and classification performance metrics of the two feature subsets across different models.

We then explored the applicability of the proposed framework in another CRGNB species. Feature selection and model development were re-executed in the CRPA cohort from the MIMIC-IV dataset, which included 286 patients treated with systemic antibiotics, including 206 survivors and 80 non-survivors. Variable definitions were aligned with those used in the development cohort. Laboratory variables were defined as the worst values from 2 days before to 2 days after CRPA positivity. Given the substantial variability of SVM and KNN in preliminary external testing, the feature-selection procedure was restricted to five models: XGBoost, CatBoost, LightGBM, random forest and logistic regression. This process yielded two feature subsets. One preferentially supported discriminative performance, including precision, AUROC and F1-score. The other favored sensitivity and overall classification performance, including accuracy, recall and AUPRC. The optimized feature subsets and their constituent variables are summarized in Supplementary Table 3. Compared with FFMs, SFMs showed similar accuracy and precision, but better recall, F1-score, AUROC and AUPRC, with significant differences (Figure 5c, all P<0.05). At the individual model level, the discrimination-oriented subset performed particularly well in tree-based and ensemble models, including an AUROC of 0.736 ± 0.060 for LightGBM. The sensitivity-oriented subset also achieved strong risk discrimination across several models, including an AUROC of 0.744 ± 0.079 for logistic regression (Figure 5d, Supplementary Figure 8). These results show that our feature-set selection framework remained able to recover stable and predictive feature subsets in a related CRGNB infection setting.

Taken together, the external analyses of the MIMIC-IV dataset provide support at two levels. First, feature subsets derived from the development cohort were transferable to an independent CRAB population. Second, the feature-set selection workflow was able to reconstruct stable and predictive feature subsets in a related, but non-identical, CRPA population. These findings suggest that this framework is not tied to a single center or a single dataset, but has broader potential as a generalizable foundation for a clinical agent in CRGNB anti-infective management.

### An interactive clinical agent operationalizes our framework for personalized decision-support

To lower the technical threshold for clinical adoption and empower practitioners lacking specialized programming or data modeling expertise, this study encapsulated the validated framework capabilities into Dr.BUG, a locally deployable, dialogue-driven clinical agent for personalized anti-infective decision support in CRGNB infection (Figure 1–2). Dr.BUG provides an executable clinical interface for the framework. It uses the large language model (LLM), Qwen-plus[51, 52], as a natural-language interface and workflow coordinator, translating clinician requests into structured backend workflows for model development, patient-level prediction and individualized treatment recommendation. This design enables clinicians to use the framework without writing code or switching between separate analytical modules, thereby improving its accessibility for routine clinical use.

In practice, Dr.BUG can connect model development with bedside decision support, as demonstrated in Supplementary Video 1. Clinicians can use local data to complete data quality review, task configuration, feature-set selection, model training, performance evaluation and versioned model release through Dr.BUG. After deployment, healthcare staff can input a patient’s clinical features together with the empirical antibiotic regimen, and the system can generate multidimensional predictions at an early stage of CRAB infection to assess the appropriateness of the current treatment strategy. Dr.BUG also returns SHAP-based explanations to identify the key factors driving the prediction. In addition, by referencing a predefined library of candidate regimens, it can generate a ranked list of alternative recommended antibiotic options to support individualized treatment decisions. The system therefore functions not as an isolated prediction tool, but as a coherent clinical workflow that links model updating with patient-specific decision support.

To preserve data privacy, all patient-level computations are executed by local modules, whereas the large language model is used primarily for request understanding, workflow coordination and result presentation. Overall, this interactive implementation translates the validated framework into a deployable clinical prototype and provides a practical technical foundation for precision anti-infective management and antimicrobial stewardship.

## Discussion

The clinical management of CRGNB infections remains challenging due to marked patient heterogeneity, limited therapeutic options, and the critical need to balance efficacy against toxicity. We developed Dr.BUG, a framework that integrates model development, prediction, and individualized regimen recommendation, lowering the operational barrier for clinicians who may not have programming or modelling expertise. Clinicians can therefore train models using datasets collected at different clinical time windows, such as before treatment or after several days of antibiotic exposure, and apply the resulting models to generate dynamic, stage-specific predictions as patient status and treatment exposure evolve. Unlike traditional risk scores that provide static prognosis, our framework extends its utility to support specific therapeutic trade-offs, offering clinicians a tool to compare candidate regimens beyond empirical intuition.

The clinical implications of this framework are multifaceted, particularly in the context of antimicrobial stewardship (AMS) and precision medicine. First, by providing individualized regimen rankings, Dr.BUG serves as a practical decision-support tool that aligns with AMS goals. It helps clinicians identify the most effective regimen while minimizing unnecessary exposure to broad-spectrum antibiotics, thereby potentially reducing selective pressure for resistance. Second, the framework facilitates precision anti-infective therapy by quantifying the risk-benefit profile of specific drug combinations for individual patients. For CRGNB infections, where “one-size-fits-all” empiric therapy is often suboptimal, this patient-specific approach provides a structured basis for optimizing drug selection, potentially improving survival rates in a population with limited therapeutic options. The stability of survival prediction across temporal and external cohorts further reinforces the reliability of this guidance, suggesting that the model captures reproducible clinical patterns rather than center-specific noise.

Methodologically, our stable feature-set selection strategy addressed a key bottleneck in translating models to clinical practice. Conventional feature selection often identifies dataset-specific “local optima” that fail to generalize. By prioritizing feature sets that perform consistently across models and partitions, our approach ensures that the input variables represent reproducible clinical signals. This robustness was confirmed through temporal and external validation in the MIMIC-IV dataset, demonstrating that the identified features are transferable across different patient populations and electronic health record systems.

Despite these contributions, several limitations should be interpreted in the context of the study design and the inherent challenges of the field. First, the development cohort size was modest, reflecting the difficulty of recruiting large cohorts of critically ill patients with rare CRGNB infections in a single-center setting. However, this limitation was actively mitigated by external validation in the multicenter MIMIC-IV dataset, which confirmed the generalizability of our findings. Second, the current models rely on static baseline variables. It is important to note that modeling dynamic, longitudinal clinical trajectories remain a recognized challenge across the field of clinical AI, and our static model represents a robust first step for initial decision support at the time of infection onset. Finally, while SHAP explanations provide interpretability, they reflect associations rather than causality; thus, the framework is intended to assist, not replace, clinical judgement.

Looking ahead, we plan to continuously refine and optimize the framework to address a broader spectrum of clinical needs in infectious disease management. Future iterations will aim to extend the applicability of Dr.BUG beyond CRGNB to cover diverse pathogen profiles and infection types. In parallel, although the present expert review provided an initial assessment of the clinical acceptability of model-prioritized regimens in patients who died, future studies should extend this evaluation to survivors, de-escalation scenarios and real-world workflows, while assessing concordance with clinician judgement, prescribing behavior and patient outcomes. With the incorporation of richer multimodal data, including pathogen genomics, resistance mechanisms, pharmacokinetic and pharmacodynamic (PK/PD) parameters, longitudinal monitoring data and unstructured clinical records, the system will evolve from a static prediction tool into a comprehensive, dynamic decision-support platform. This evolution will enable the framework to support clinicians throughout the entire patient journey, further advancing the realization of precision anti-infective therapy.

In conclusion, Dr.BUG operationalizes a validated framework into a deployable clinical agent. By bridging the gap between complex predictive analytics and bedside decision-making, it shifts the role of AI from passive prognostication to active therapeutic support. With responsible integration into practice, such systems hold the potential to standardize CRGNB management, optimize antibiotic use, and ultimately improve patient outcomes in this era of increasing antimicrobial resistance.

## Methods

### Study population

The development and temporal validation cohorts were derived from Ruijin Hospital, Shanghai Jiao Tong University School of Medicine, China. The study population comprised hospitalized patients with microbiologically confirmed CRAB infection who received systemic antibiotic therapy.

Patients were eligible for inclusion if they met the following criteria: (1) microbiological confirmation of CRAB infection; (2) receipt of polymyxin-based therapy; and (3) availability of complete baseline clinical data and key laboratory measurements. Only the first infection episode for each patient was retained. Patients with missing key outcome data were excluded. Between October 2020 and September 2024, 443 patients met the eligibility criteria. Of these, 2 were excluded because of missing outcome data, yielding a final development cohort of 441 patients. This cohort was used for feature-set selection, model development, and internal evaluation.

In addition, an independent temporal validation cohort from the same center and two external cohorts derived from the MIMIC-IV dataset (v3.1)[50] were constructed for validation. The temporal validation cohort was used to assess model robustness under temporal distribution shift. The external cohorts were used to evaluate the cross-center generalizability of the selected feature sets and the transferability of the feature-set selection framework.

### Task definitions

To capture multiple clinically relevant dimensions of CRAB management, we formulated a multitask prediction framework comprising three binary classification tasks and one regression task: clinical efficacy, survival outcome, polymyxin resistance and antibiotic treatment duration.

Clinical efficacy prediction was defined as a binary endpoint. A positive outcome indicated clinical improvement within 21 days of treatment, whereas a negative outcome indicated treatment failure. Clinical improvement was determined using an integrated clinical assessment, including sustained normalization of body temperature, improvement in oxygenation status, reduced respiratory rate, marked decreases in inflammatory markers (white blood cell count, C-reactive protein and procalcitonin), and radiological stabilization or resolution. Patients who did not meet these criteria or showed persistent clinical deterioration were classified as treatment failure.

Survival outcome prediction was defined according to in-hospital status. Survival to hospital discharge was labeled as positive, whereas in-hospital death or discharge because of clinical deterioration was labeled as negative. This task was intended to capture overall survival during hospitalization.

Polymyxin resistance prediction was formulated as a binary classification task for early identification of resistance risk. The task definition was based on antimicrobial susceptibility testing results. It was developed with reference to the CLSI M100 performance standards[53] and further refined through discussion with clinical experts. In this study, isolates with a polymyxin MIC of ≥2 μg/mL were operationally classified as resistant or resistance-risk positive. Isolates with a polymyxin MIC of <2 μg/mL were classified as susceptible or resistance-risk negative.

Antibiotic treatment duration prediction was formulated as a regression task to estimate the duration of antibiotic therapy for each patient.

### Variables and preprocessing

Candidate features were drawn from seven domains: demographics, comorbidities, immunosuppression status, infection-related clinical conditions, laboratory measurements, microbiological findings, and antimicrobial exposure with susceptibility profiles. To minimize information leakage, feature construction was restricted to data available within an early time window around infection diagnosis and treatment initiation. Variables with more than 30% missingness were excluded, and the remaining variables underwent standardized preprocessing.

Demographic variables included age, sex, height, weight and body mass index (BMI). Comorbidities and immunosuppression status were encoded as binary indicator variables, with detailed definitions provided in Supplementary Table 1. Infection-related clinical variables included infection type, respiratory support modality, inspired oxygen concentration and receipt of renal replacement therapy. Infection type was categorized as ventilator-associated pneumonia (VAP) or hospital-acquired pneumonia (HAP), and respiratory support was encoded as an ordinal variable reflecting increasing levels of support intensity.

Laboratory variables captured inflammatory, hematological, coagulation, renal and hepatic status. White blood cell count, neutrophil percentage, absolute lymphocyte count, platelet count, C-reactive protein (CRP), procalcitonin (PCT) and D-dimer were defined as the most adverse values within a 2-day window before and after CRAB diagnosis, according to the direction of clinical deterioration. Baseline creatinine, estimated glomerular filtration rate (eGFR), alanine aminotransferase (ALT), aspartate aminotransferase (AST), total bilirubin and serum albumin were defined as the most recent measurements obtained before initiation of antimicrobial therapy.

Microbiological variables were used to characterize co-infecting pathogens on the basis of microbiological findings obtained within 2 days before and after CRAB identification, integrating both conventional culture and next-generation sequencing data. These findings were further categorized as Gram-negative bacteria, Gram-positive bacteria or fungi.

Antimicrobial exposure variables were used to characterize treatment regimens. Polymyxin exposure was represented by daily administration frequency, whereas concomitant antibiotics were quantified by average daily dose. Antimicrobial susceptibility results were classified as resistant, susceptible or intermediate according to clinical standards, with resistant and intermediate categories grouped as non-susceptible. Susceptibility variables with more than 30% missingness were excluded. For retained variables, missing susceptibility results were conservatively coded as susceptible.

During preprocessing, missing values in continuous variables were imputed using the median, and missing values in categorical variables using the mode. Feature scaling was applied according to model class: tree-based models were trained without standardization, whereas linear and distance-based models were standardized before training. To avoid information leakage during model evaluation, imputation and scaling were performed within each training fold and then applied to the corresponding validation fold. For classification tasks with class imbalance, built-in balancing strategies were used when supported by the model.

### Consensus-driven hierarchical feature set selection strategy

To improve model interpretability while enhancing the robustness and transferability of selected feature subsets across model families and datasets, we developed a consensus-driven hierarchical feature-set selection strategy. This procedure integrated SHAP-based feature attribution, exhaustive subset enumeration and cross-model consensus evaluation to identify stable and high-performing feature combinations.

We first defined seven model families, namely Extreme Gradient Boosting (XGBoost), Light Gradient Boosting Machine (LightGBM), Categorical Boosting (CatBoost), random forest (RF), logistic regression (LR), support vector machine (SVM) and k-nearest neighbors (KNN). For each prediction task, the corresponding classification or regression variants were fitted in the full feature space using five-fold cross-validation under default hyperparameters. SHAP values[45] were computed within each fold to quantify feature contributions, using TreeSHAP for tree-based models, LinearSHAP for linear models and KernelSHAP for the remaining models. For each model, global feature importance was defined as the mean absolute SHAP value averaged across folds, yielding a model-specific ranking.

To account for the structural properties of clinical variables, we applied a hierarchical attribution scheme. Antibiotic-related variables were treated as baseline intervention features. In tasks directly related to treatment response and prognosis, these variables were excluded from competitive ranking but appended to all candidate subsets during subsequent model fitting. For sparse categorical modules, such as comorbidities and immunosuppression status, which were represented by multiple one-hot encoded variables, feature importance was aggregated at the module level to avoid fragmentation. Specifically, the importance of feature group *g* was defined as:

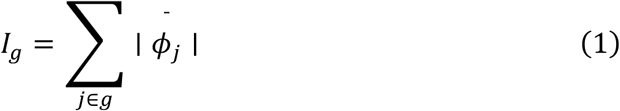

where ∣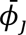∣ denotes the mean absolute SHAP value of feature *j* across folds.

Following baseline exclusion and module aggregation, the top 10 ranked features from each model were retained to form model-specific candidate pools.

To identify informative feature combinations beyond marginal effects, exhaustive subset enumeration was performed within each candidate pool. All possible subsets with sizes ranging from 1 to 10 were generated. This strategy was used to capture potential non-linear interactions and complementary effects among variables, because subsets composed of individually lower-ranked features may outperform those formed solely from the highest-ranked features. During model fitting, antibiotic-related variables, when predefined as mandatory for a given task, were appended to each candidate subset, and grouped variables were expanded back to their original one-hot encoded representations.

Each candidate subset was then evaluated by five-fold cross-validation. For classification tasks, subset performance was quantified by mean AUROC:

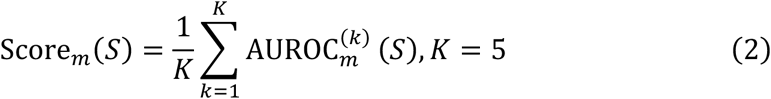

For the regression task, subset performance was quantified by mean Pearson correlation coefficient (PCC):

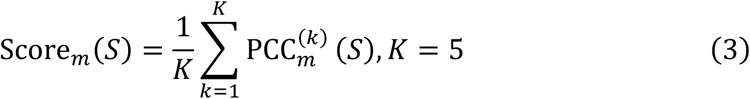

where *S* denotes a candidate feature subset and *m* denotes the model. For each model, the top seven subsets were retained, yielding a pooled set of up to 49 candidate feature subsets for each task.

We next performed cross-model consensus evaluation across the pooled candidate subsets. Each retained subset was re-evaluated using all seven model families with five-fold cross-validation. For classification tasks, performance was assessed using Accuracy, Precision, Recall, F1-score, AUROC and AUPRC. For the regression task, performance was assessed using PCC. To reduce the influence of model-specific outlier subsets, low-ranking subsets across multiple models were excluded according to a predefined filtering rule before cross-model aggregation. Consensus performance under metric *q* was then calculated as:

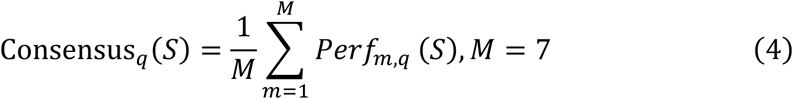

where *Per_m,q_*(*S*) denotes the mean cross-validated performance of subset *S*under metric *q* in model *m*. Final feature subsets were selected from the retained candidates according to their relative consensus performance. When appropriate, different subsets were retained for different target metrics.

This two-stage selection framework was designed to balance predictive performance, parsimony and cross-model stability, thereby improving the robustness and transferability of the selected feature subsets across algorithms and datasets.

### Model development and evaluation

To assess the robustness of the selected feature sets across different modelling frameworks, we evaluated seven model families: XGBoost, LightGBM, CatBoost, RF, LR, SVM and KNN. For each task, the corresponding classification or regression variants were trained and internally evaluated in the development cohort using five-fold cross-validation. Unless otherwise specified, the same data partitioning and preprocessing procedures were applied across algorithms, and models were trained using standard parameter settings.

For classification tasks, we employed Accuracy, Precision, Recall, F1-score, the area under the receiver operating characteristic curve (AUROC), and the area under the precision-recall curve (AUPRC) to assess model performance. These metrics are defined as follows:

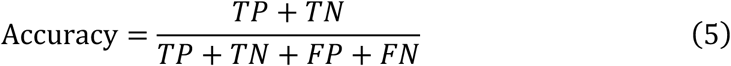

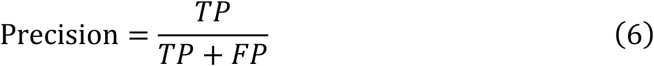

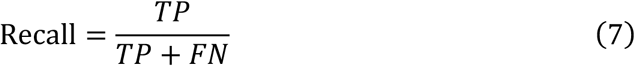

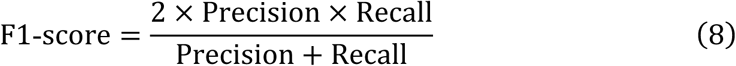

where *TP*, *TN*, *FP*, and *FN* denote true positives, true negatives, false positives, and false negatives, respectively. AUROC measures overall discriminative ability across decision thresholds, whereas AUPRC provides a complementary evaluation under class imbalance by emphasizing performance on the positive class.

For the regression task, performance was evaluated using both mean squared error (MSE) and the Pearson correlation coefficient (PCC). These metrics are defined as follows:

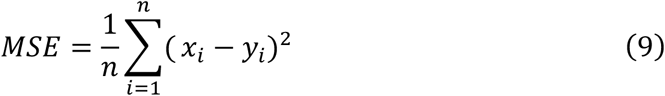

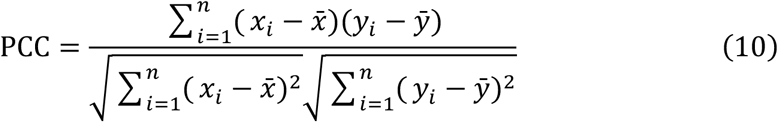

where *x_i_* and *y_i_* denote the observed and predicted values, respectively, and *x̅* and *y̅* denote their means.

For classification tasks, performance differences between feature strategies were assessed using paired one-sided Wilcoxon signed-rank tests. For each task, performance metrics obtained from the FFMs and the corresponding SFMs across model-fold evaluations were treated as paired observations, allowing an integrated comparison of the overall effect of feature reduction across modelling frameworks. Statistical significance was denoted as ∗ *P* < 0.05, ∗∗ *P* < 0.01, and ∗∗∗ *P* < 0.001. In addition, we calculated a metric-aligned matched-or-exceeded rate to summarize how often SFMs preserved or improved upon FFM performance. For each selected feature set, comparisons were restricted to the metric or metrics used to derive that feature set. For example, if a survival feature set was optimized for precision and AUROC, the corresponding SFM was compared with its FFM counterpart only on precision and AUROC. A comparison was counted as matched or exceeded when the SFM achieved a value equal to or better than the corresponding FFM for that metric. For metrics in which higher values indicate better performance, including accuracy, precision, recall, F1-score, AUROC and AUPRC, equality or a higher value was counted. For MSE, equality or a lower value was counted. The overall matched-or-exceeded rate was calculated as the number of matched-or-exceeded comparisons divided by the total number of metric-aligned comparisons.

### SHAP-based interpretability analysis

To improve model interpretability, SHAP (Shapley Additive Explanations)[45] was used to explain model predictions. SHAP is a game-theoretic framework derived from Shapley values that decomposes the contribution of each feature to the model output in an additive manner, thereby quantifying the relative contribution of individual variables to prediction. These analyses were used to characterize the basis of model predictions and patterns of variable contribution, rather than to infer causal relationships between predictors and outcomes.

For global interpretability, SHAP values were calculated for trained models, and overall feature importance was quantified using the mean absolute SHAP value of each variable to rank the principal drivers of prediction. For categorical variables represented by multiple subcategories, such as comorbidities and immunosuppression status, one-hot encoded components derived from the same original variable were aggregated before visualization to improve interpretability.

For local interpretability, patient-level SHAP values were computed for individual predictions generated by the SFMs. Waterfall plots or force plots were used to visualize the direction and magnitude of each variable’s contribution to the predicted outcome. In addition, for tree-based models, SHAP interaction values were calculated to characterize relationships among input features, and feature interaction networks were constructed accordingly. Variables included in the interaction analysis comprised both the selected clinical features and medication-related features. Categorical variables with multiple subcategories were likewise aggregated to the level of the original variable for presentation.

### Temporal validation

To assess temporal robustness, an independent temporal validation cohort was constructed by consecutively enrolling new patients from the same center between October 2024 and May 2025, using the same inclusion and exclusion criteria as those applied to the development cohort. In total, 111 patients were included.

Models developed in the development phase were directly applied to the temporal validation cohort without retraining. Both model parameters and selected feature sets were fixed, and no additional feature selection or model updating was performed. Performance was evaluated using the same metrics as in the development cohort to assess generalizability under temporal distribution shift.

### External validation

The MIMIC-IV database (v3.1) was used for external validation. MIMIC-IV is a large, publicly available de-identified electronic health record database containing hospital data from Beth Israel Deaconess Medical Center in Boston, Massachusetts, USA, between 2008 and 2019. Two external cohorts were constructed for distinct validation purposes.

To evaluate the cross-center generalizability of the selected feature sets, an external CRAB cohort was constructed from the MIMIC-IV dataset. Patients were included if they had at least one positive culture for *A. baumannii* during hospitalization, documented resistance to at least one carbapenem, and receipt of systemic antibiotic therapy. Only the first infection episode for each patient was retained, and patients with missing outcome data were excluded. A total of 42 patients were included, of whom 9 died in hospital or were transitioned to end-of-life care and 33 survived. This cohort was used to externally validate the feature sets identified for the survival prediction task in the development cohort. Because of differences in variable availability in the MIMIC-IV dataset, only variables included in the selected feature sets, together with antibiotic exposure variables encoded consistently with the development cohort, were extracted. The feature sets were transferred directly without re-running the selection procedure. Given the limited sample size, model performance was evaluated using leave-one-out cross-validation (LOOCV).

To assess the transferability of the feature-set selection framework, we further constructed an external CRPA cohort from the MIMIC-IV dataset. Adult patients (aged ≥18 years) with at least one positive culture for *Pseudomonas aeruginosa* showing resistance to at least one carbapenem were included. For patients with multiple positive cultures during a single hospitalization, only the first culture was retained as the index event. For patients with multiple hospital admissions, only the first admission with a CRPA event was included. Patients were excluded if key microbiological data were missing, if 28-day all-cause mortality status could not be ascertained, or if antibiotic therapy consisted of a single administration or lasted for ≤24 hours. A total of 286 patients were included, comprising 80 non-survivors and 206 survivors within 28 days. This cohort was not used for direct transfer of fixed feature sets. Instead, the feature-set selection workflow was re-run in this dataset, and the newly derived feature sets were then used for model development. Performance of models based on selected feature sets was compared with that of FFMs for 28-day all-cause mortality prediction, in order to assess the transferability of the feature-set selection framework.

### Individualized antibiotic recommendation simulation

Individualized antibiotic recommendation simulation was performed for the survival prediction task. A survival prediction model was first trained in the development cohort using the selected feature set. The trained model was then fixed and applied to the temporal validation cohort, in which candidate antibiotic regimens were enumerated and re-scored at the patient level.

Candidate antibiotic regimens were defined according to drug composition and dosing intensity. Each regimen was encoded using drug-related variables, with polymyxin-based treatment represented by administration frequency and concomitant antibiotics encoded according to daily dose. The candidate regimen space included monotherapy, dual therapy and triple-drug combinations, and all eligible combinations were generated under predefined constraints.

For each patient, non-antibiotic clinical features were held constant, whereas drug-related variables were replaced by the encoding of each candidate regimen. The modified feature vectors were then entered into the trained survival model to estimate the corresponding predicted survival probabilities. Candidate regimens were ranked according to predicted survival probability, and the top-ranked regimen was defined as the model-prioritized regimen for that patient.

### Implementation of Dr.BUG

Dr.BUG was implemented using a decoupled front-end/back-end architecture. The back end, built on FastAPI, handled dialogue requests, task orchestration, model invocation and result aggregation. The front end, developed with Vue and Vite, provided an interactive interface for user input, parameter configuration, task monitoring and result visualization. A model registry and task management system were incorporated to maintain versioned model metadata and feature schemas, and to support training job submission, model release and prediction result management.

Qwen-Plus[51, 52] was integrated as the conversational and orchestration layer for request interpretation, read-only tool planning, context-aware response generation and result presentation. Upon receiving a natural-language request, the system parsed the user intent together with the current task context and routed the corresponding structured task to local deterministic modules for feature processing, model training, patient-level prediction, antibiotic-regimen rescoring and interpretability analysis. Structured outputs were then generated and returned through the interactive interface. Model training, patient-level prediction and regimen rescoring were performed by locally deployed deterministic modules.

To reduce unnecessary exposure of sensitive clinical data, the language model did not directly access raw patient-level tabular data, but interacted with the system through constrained tools that exposed only privacy-filtered, task-relevant information.

## Ethics

The study was conducted in accordance with the Declaration of Helsinki and relevant institutional guidelines. The study was approved by the Ruijin Hospital Ethics Committee, Shanghai Jiao Tong University School of Medicine. The requirement for informed consent was waived by the Ethics Committee due to the retrospective nature of the study.

The MIMIC-IV dataset used for external validation is a publicly available, de-identified electronic health record database hosted on PhysioNet. The collection of patient information and the creation of the research resource were reviewed by the Institutional Review Board of Beth Israel Deaconess Medical Center, which granted a waiver of informed consent and approved the data-sharing initiative. Access to MIMIC-IV was obtained after completion of the required training and data use agreement.

## Data Availability

The MIMIC-IV database used for external validation in this study is available through the PhysioNet repository after completion of the required credentialing process and data use agreement. The development cohort and temporal validation cohort were derived from institutional clinical records and cannot be made publicly available because of regulatory, ethical and privacy restrictions. Access to de-identified data from these cohorts may be considered by the corresponding author upon reasonable request, subject to institutional review, ethics approval where required, and execution of an appropriate data use agreement.

## Code Availability

The source code of Dr.BUG is available at https://github.com/cccccxw0909/Dr.BUG. A demonstration video illustrating the main workflow of Dr.BUG is available on figshare at https://doi.org/10.6084/m9.figshare.32221362.

