## Supplementary Information for "An interpretable and interactive clinical AI agent for personalized anti-infective decision support in carbapenem-resistant Gram-negative bacterial infection"

**Supplementary Note 1. Feature subsets improve discriminative performance and robustness in clinical efficacy prediction task**

In the clinical efficacy prediction task, four representative feature subsets were identified, each optimized for a different evaluation objective. Overall, the SFMs retained performance comparable to that of the FFMs for Accuracy, Recall, F1-score and AUPRC, indicating that most predictive information was preserved despite the reduction in input variables (Supplementary Figure 2a). The clearest benefit was observed in discriminative performance. Precision and AUROC both showed an upward shift in their cross-validation distributions, and both improvements reached statistical significance (Supplementary Figure 2a; Precision P<0.001, AUROC P<0.01). This pattern was also reflected in the ROC analyses, in which the selected feature subsets generally maintained, and in several models modestly improved, AUC relative to the corresponding FFMs (Supplementary Figure 3). Using Random Forest as a representative example, feature-set A achieved an Accuracy of 0.760, a Precision of 0.762, a Recall of 0.982, an F1-score of 0.858, an AUROC of 0.710 and an AUPRC of 0.868, compared with 0.760, 0.758, 0.994, 0.860, 0.682 and 0.822, respectively, for the FFM (Supplementary Figure 2b-c). Unlike the survival task, the advantage of feature selection in this setting was not a uniform increase across all metrics. Instead, it was expressed as improved discrimination with little loss of overall predictive capacity. These results suggest that, for clinical efficacy prediction, the key signal can also be captured by a smaller set of more informative variables, which may reduce redundancy and improve the robustness of model discrimination.

**Supplementary Note 2. Feature subsets improve predictive performance and algorithmic robustness in** **polymyxin resistance prediction task**

Polymyxin resistance prediction showed a broader and more consistent benefit from feature selection. Five representative feature subsets were identified. Compared with the FFMs, the SFMs significantly improved Precision, Recall, F1-score, AUROC and AUPRC, while maintaining comparable Accuracy (Supplementary Figure 4; all five metrics, P<0.001). This advantage was not limited to isolated models. CatBoost, LightGBM, XGBoost, KNN and SVM all showed overall performance gains after restriction to the optimized feature subsets. The ROC curves further supported this pattern, with most representative subsets showing improved or more stable discrimination than the corresponding FFMs across models (Supplementary Figure 5). Together, these findings indicate that polymyxin resistance is more effectively captured by a compact set of highly discriminative variables than by the full input space. In this task, feature selection improved predictive performance and enhanced algorithmic robustness in a more consistent manner.

**Supplementary Note 3. Clinically coherent physiological signals underlie treatment response prediction**

For clinical efficacy prediction, the Random Forest-FFM identified oxygen support intensity, comorbidity burden, inflammatory activity and renal function as the main contributors to model output (Supplementary Figure 6a). These core information domains were retained in the corresponding reduced feature set, indicating that the principal signals associated with treatment response could be preserved after feature reduction. In the XGBoost-SFM, higher C-reactive protein levels, greater oxygen support requirements and poorer baseline renal function were associated with a lower predicted probability of clinical improvement (Supplementary Figure 6b). Together, these features describe a clinically coherent pattern of persistent inflammation, respiratory compromise and limited physiological reserve. The interaction analysis further showed that renal function, inflammatory burden, oxygen requirement and baseline host status were tightly connected (Supplementary Figure 6c), suggesting that the model relied on an integrated clinical state rather than on isolated variables. Overall, these findings indicate that the SFM preserved the main clinically relevant signals for treatment response while remaining more parsimonious and easier to interpret.

**Supplementary Note 4. Baseline vulnerability and inflammatory disturbance contribute to polymyxin resistance prediction**

For polymyxin resistance prediction, the XGBoost-FFM assigned the highest importance to baseline patient characteristics, including age and comorbidity burden, together with infection-phase laboratory variables such as platelet count, aspartate aminotransferase and C-reactive protein (Supplementary Figure 6d). These domains were also retained in the optimized reduced feature set, indicating that the main signals relevant to resistance prediction were preserved after dimensionality reduction. In the CatBoost-SFM, older age, heavier comorbidity burden and greater inflammatory activity were associated with a higher predicted probability of polymyxin resistance (Supplementary Figure 6e). This pattern suggests that resistance prediction was linked not to a single abnormal marker, but to the combined influence of baseline vulnerability and acute physiological disturbance. The interaction analysis further showed that baseline characteristics modified the contribution of laboratory variables and formed a relatively stable predictive structure with them (Supplementary Figure 6f). Taken together, these results indicate that the SFM retained the principal clinically relevant signals for resistance prediction while improving interpretability.

**Supplementary Note 5. Temporal validation shows partial robustness of clinical efficacy prediction**

In the clinical efficacy prediction task, temporal robustness was more moderate than that observed for survival. Performance still varied across classifiers, and some metrics showed mild attenuation after transfer to the temporal cohort. However, high sensitivity and overall classification capacity were largely preserved in several SFMs (Supplementary Figure 7b). In Random Forest, for example, multiple SFMs maintained accuracy above 0.72, while Precision, Recall, F1-score and AUPRC all remained above 0.80. By contrast, AUROC was lower, at approximately 0.55-0.62. These findings suggest that the selected feature subsets could still capture part of the signal associated with clinical improvement in temporally independent patients. However, their ability to rank patients according to outcome probability was weaker than that observed in the development cohort.

**Supplementary Note 6. Temporal validation reveals heterogeneous but clinically relevant signals in polymyxin resistance prediction**

In the polymyxin resistance prediction task, temporal performance was more heterogeneous and more strongly dependent on classifier choice. The SFMs did not produce a uniform improvement across models. However, they did improve the recognition of resistant cases in several settings (Supplementary Figure 7b). A representative example was Random Forest. In the Random Forest-FFM, the model failed to identify positive samples, with Precision, Recall and F1-score all equal to 0. After restriction to selected feature subsets, the Random Forest-SFM recovered the ability to detect a proportion of resistant cases. These findings suggest that, in a task marked by class imbalance and sparse positive signals, feature reduction may not consistently improve overall performance. Even so, it may help prevent some FFMs from collapsing on the minority class and partially restore clinically relevant detection of resistant patients.

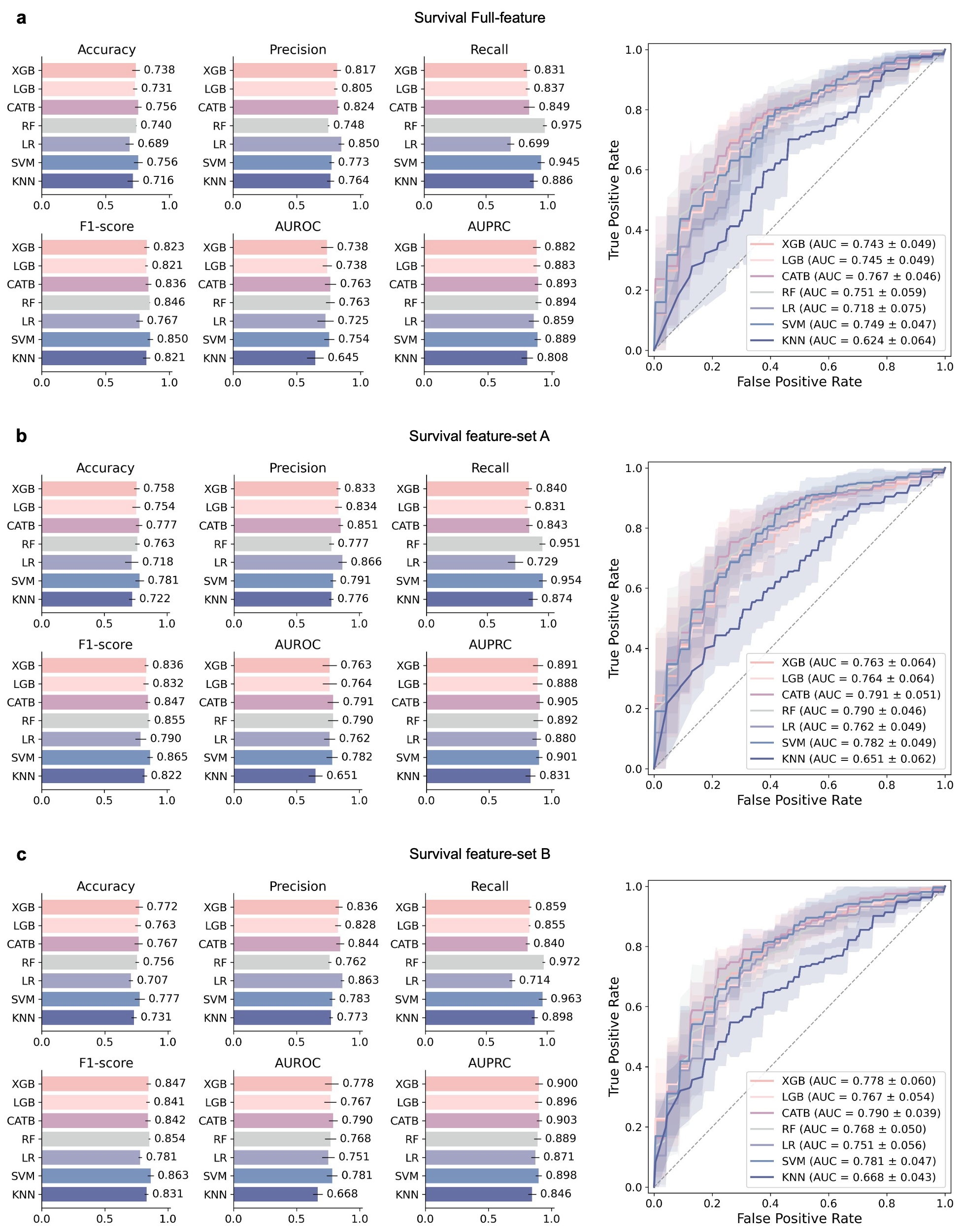

**Supplementary Figure 1. Model performance for survival prediction in the development cohort.**

(a) Overall performance of FFMs for clinical efficacy prediction.

(b-c) Overall performance of the two optimized feature subsets (feature sets A-B) across different models for survival prediction and the corresponding ROC curves; shaded areas denote standard deviation.

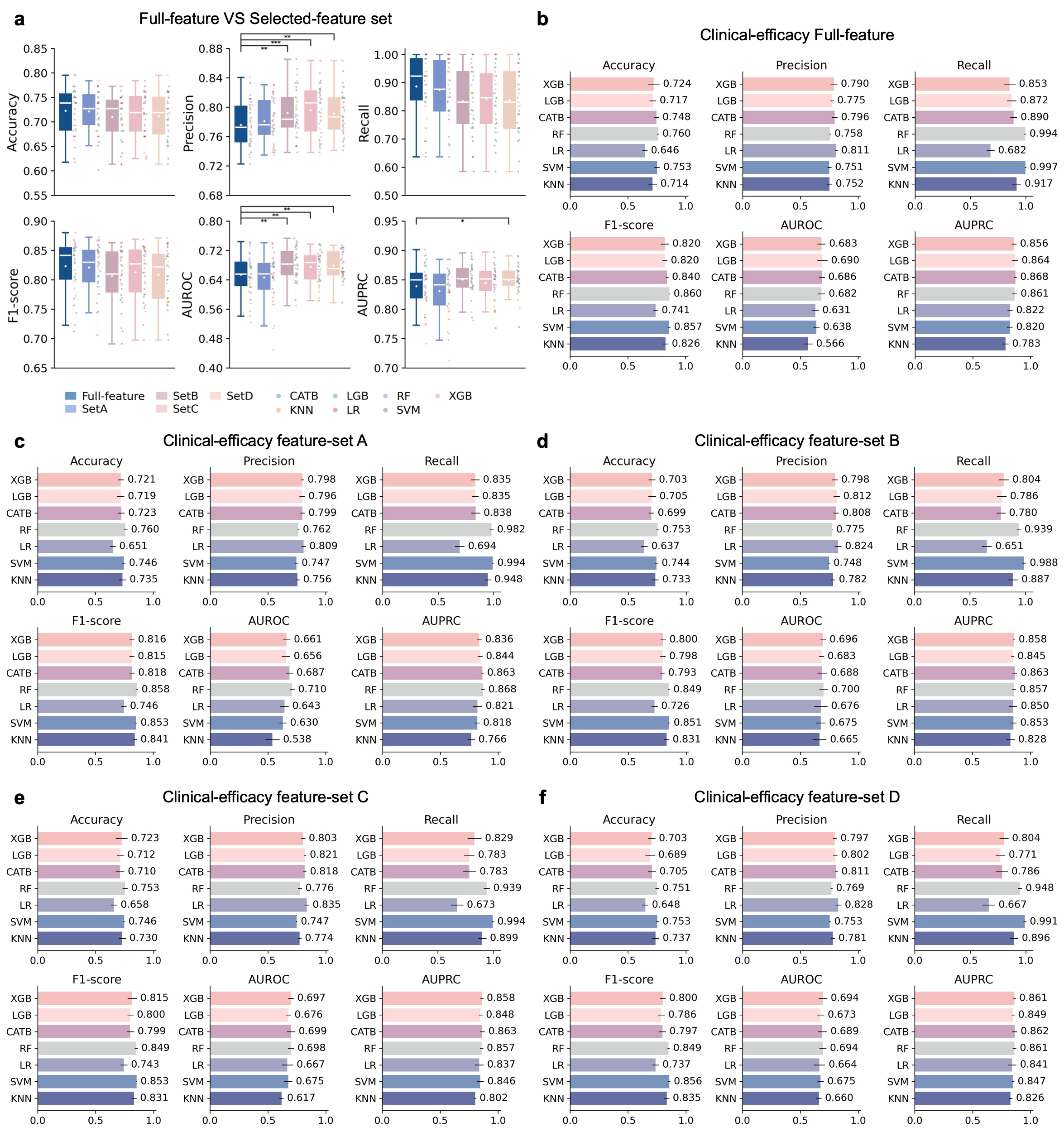

**Supplementary Figure 2. Model performance for clinical efficacy prediction in the development cohort.**

(a) Fivefold cross-validation comparison between FFMs and SFMs for clinical efficacy prediction. Dots in the boxplots indicate results from individual models across cross-validation folds; boxes represent the interquartile range and center lines denote the median. Statistical significance was assessed using one-sided paired Wilcoxon signed-rank tests: P<0.05, *P<0.01, and **P<0.001.

(b) Overall performance of FFMs for clinical efficacy prediction.

(c-d) Overall performance of four representative optimized feature subsets (feature sets A-D) across different models for clinical efficacy prediction.

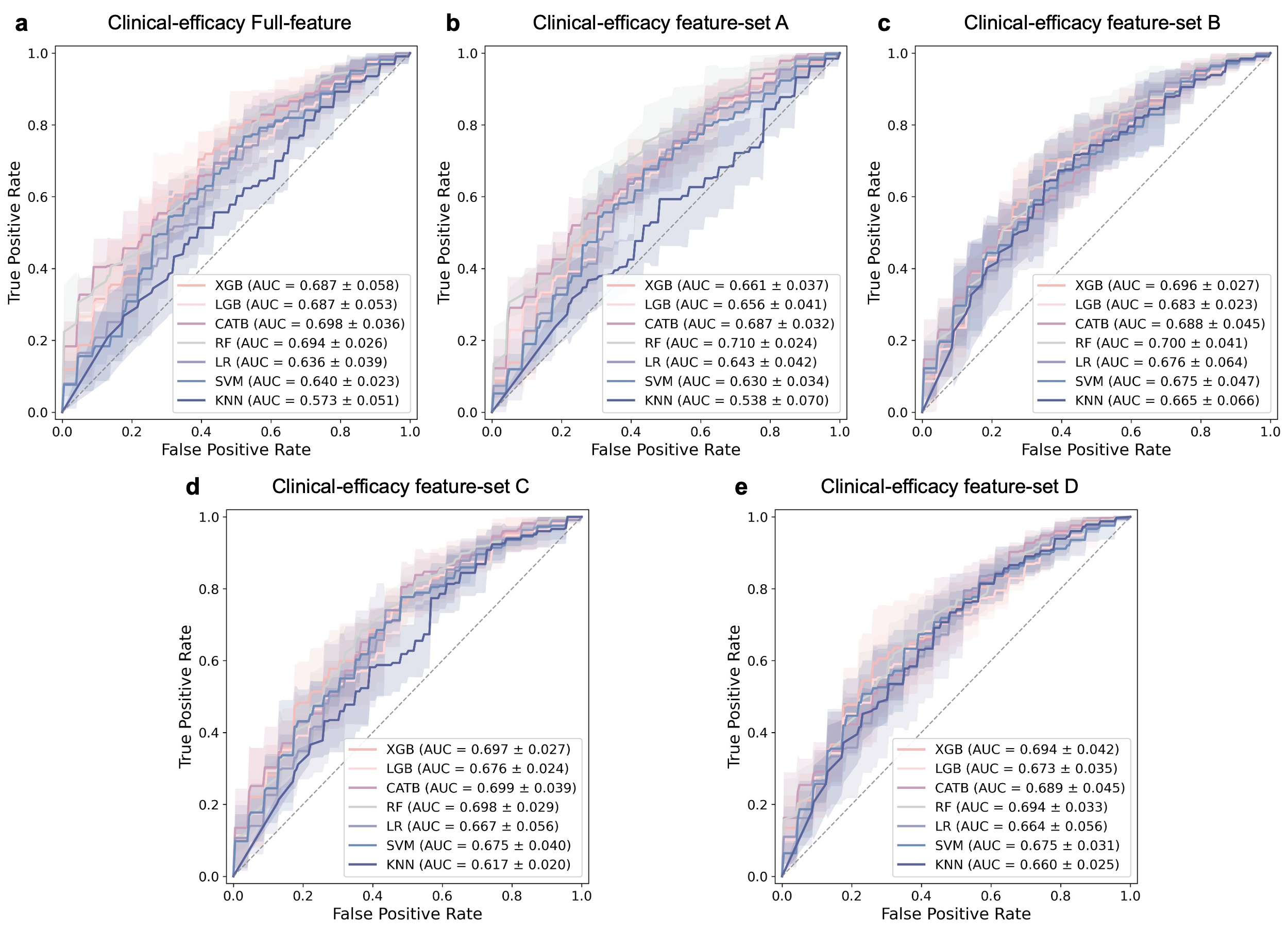

**Supplementary Figure 3. ROC curves of FFMs and representative SFMs for clinical efficacy prediction in the development cohort.**

(a) ROC curves of FFMs across different models for clinical efficacy prediction.

(b-e) ROC curves of four representative optimized feature subsets (feature sets A-D) across different models for clinical efficacy prediction. Shaded areas indicate variability across fivefold cross-validation.

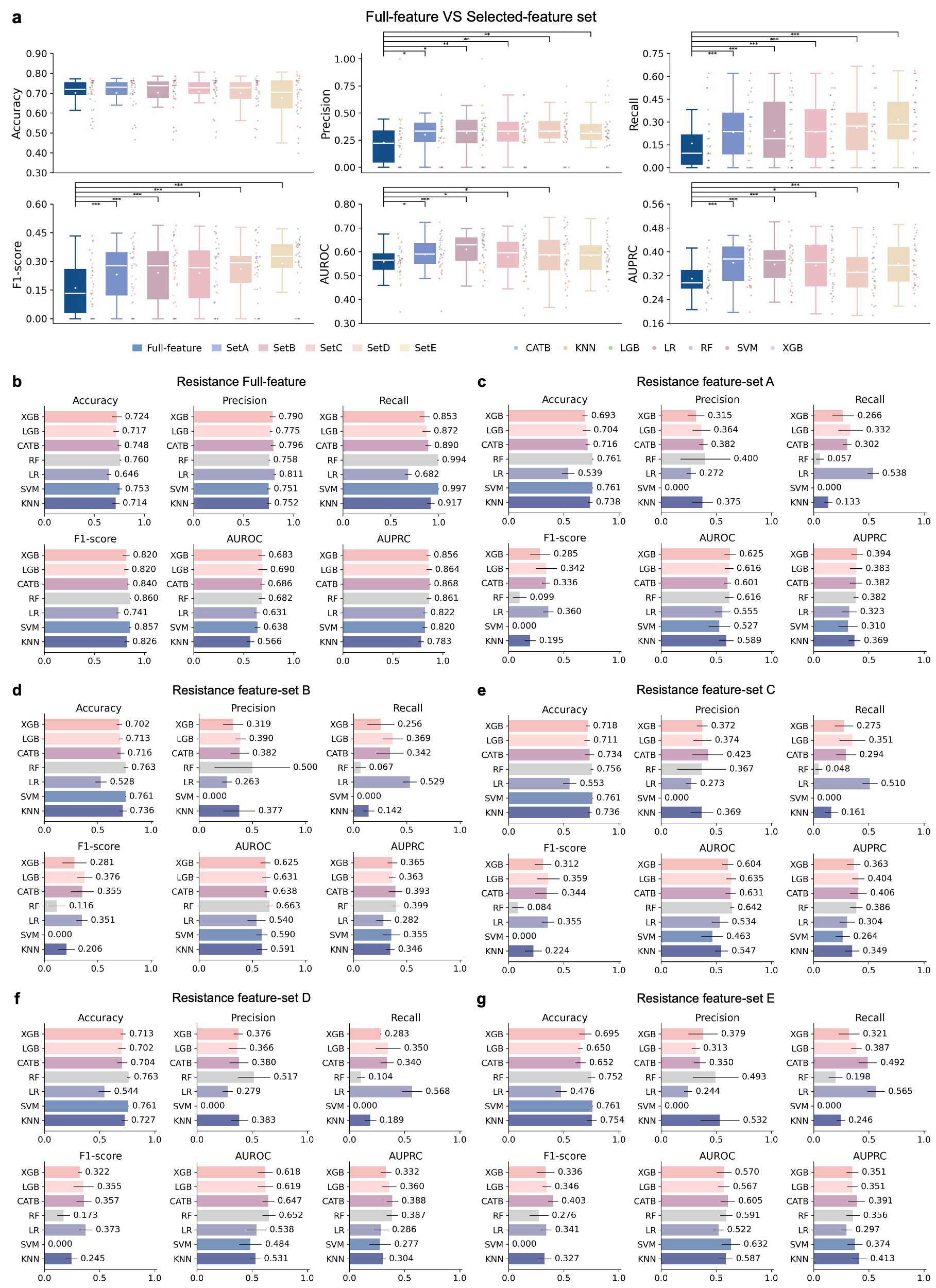

**Supplementary Figure 4. Model performance for** **polymyxin resistance prediction in the development cohort.**

(a) Fivefold cross-validation comparison between FFMs and SFMs for polymyxin resistance prediction. Dots in the boxplots indicate results from individual models across cross-validation folds; boxes represent the interquartile range and center lines denote the median. Statistical significance was assessed using one-sided paired Wilcoxon signed-rank tests: P<0.05, *P<0.01, and **P<0.001.

(b) Overall performance of FFMs for polymyxin resistance prediction.

(c-f) Overall performance of four representative optimized feature subsets (feature sets A-E) across different models for polymyxin resistance prediction.

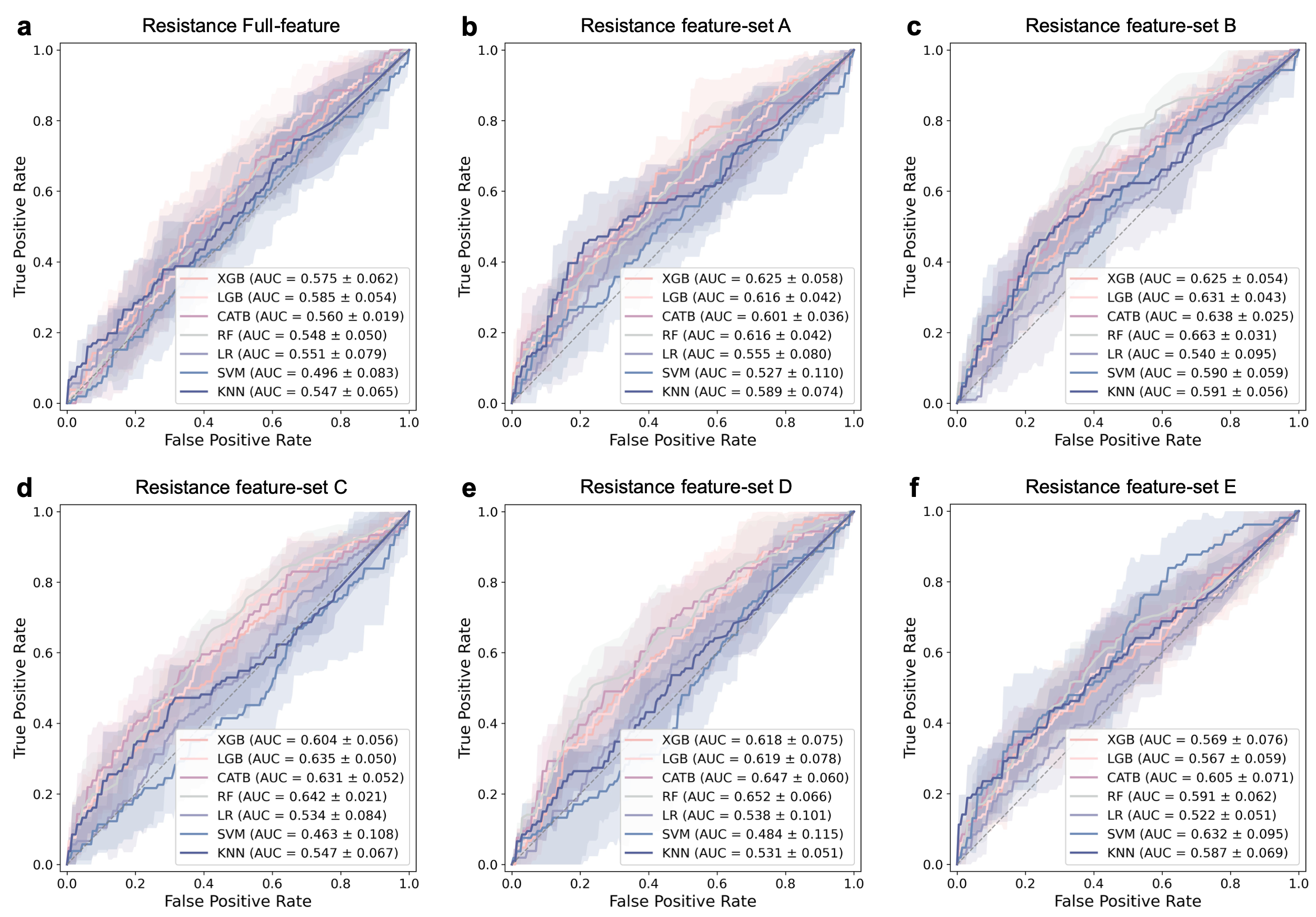

**Supplementary Figure 5. ROC curves of FFMs and representative SFMs** **for** **polymyxin resistance prediction in the development cohort.**

(a) ROC curves of FFMs across different models for polymyxin resistance prediction.

(b-f) ROC curves of four representative optimized feature subsets (feature sets A-E) across different models for polymyxin resistance prediction. Shaded areas indicate variability across fivefold cross-validation.

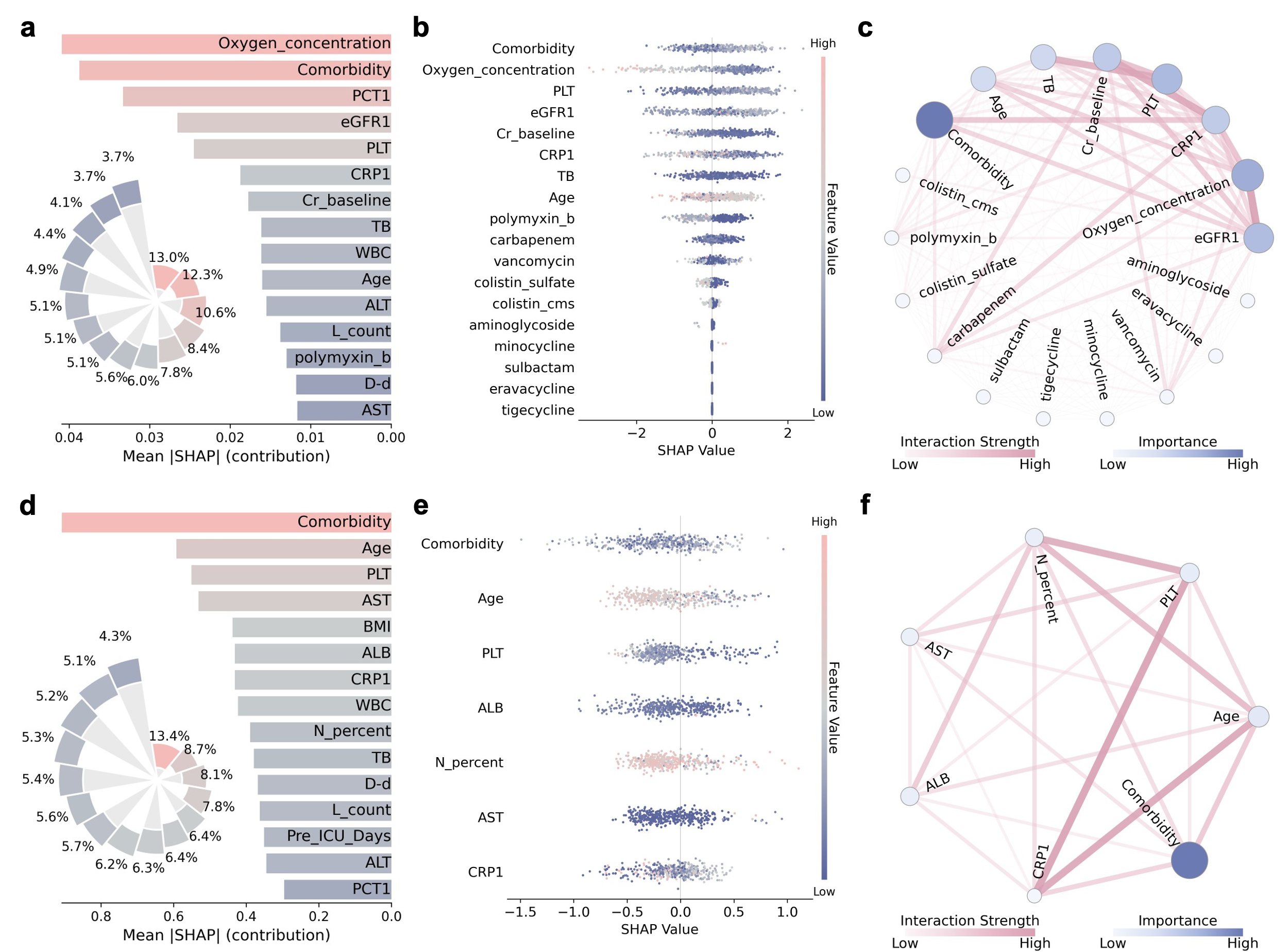

**Supplementary Figure 6. Interpretability analysis for** **clinical efficacy and** **polymyxin resistance prediction task.**

(a) Global feature importance ranking of the Random Forest-FFM for clinical efficacy, showing the top 15 features ranked by mean absolute SHAP value, and the top 10 features were retained in the candidate pool for subsequent feature-set selection.

(b) SHAP beeswarm plot of the XGBoost-SFM on clinical-efficacy feature-set A showing the direction and magnitude of feature contributions to model output.

(c) Feature interaction network of the same XGBoost-SFM.

(e) Global feature importance ranking of the XGBoost-FFM for polymyxin resistance.

(f) SHAP beeswarm plot of the CatBoost -SFM on polymyxin-resistance feature-set A. Each dot represents one patient; color indicates feature value from low (blue) to high (red), and the sign of the SHAP value indicates positive or negative contribution to the model output.

(g) Feature interaction network of the same CatBoost -SFM. Node size and color intensity indicate global feature importance, and edge width reflects interaction strength between features.

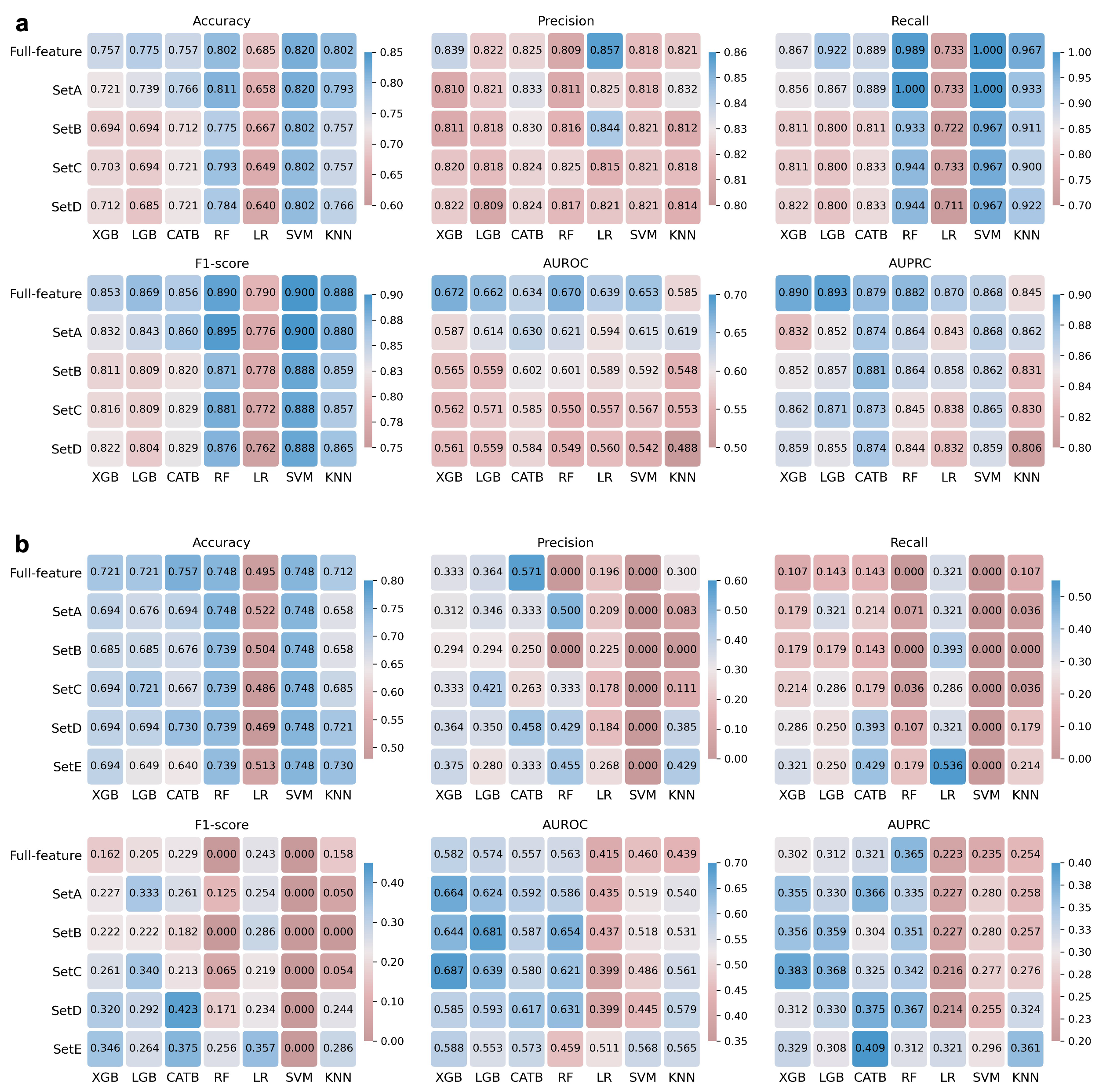

**Supplementary Figure 7. model performance for** **clinical efficacy and** **polymyxin resistance prediction in the temporal validation cohort.**

(a) Performance comparison between FFMs and SFMs for clinical efficacy in the temporal validation cohort.

(b) Performance comparison between FFMs and SFMs for polymyxin resistance prediction in the temporal validation cohort.

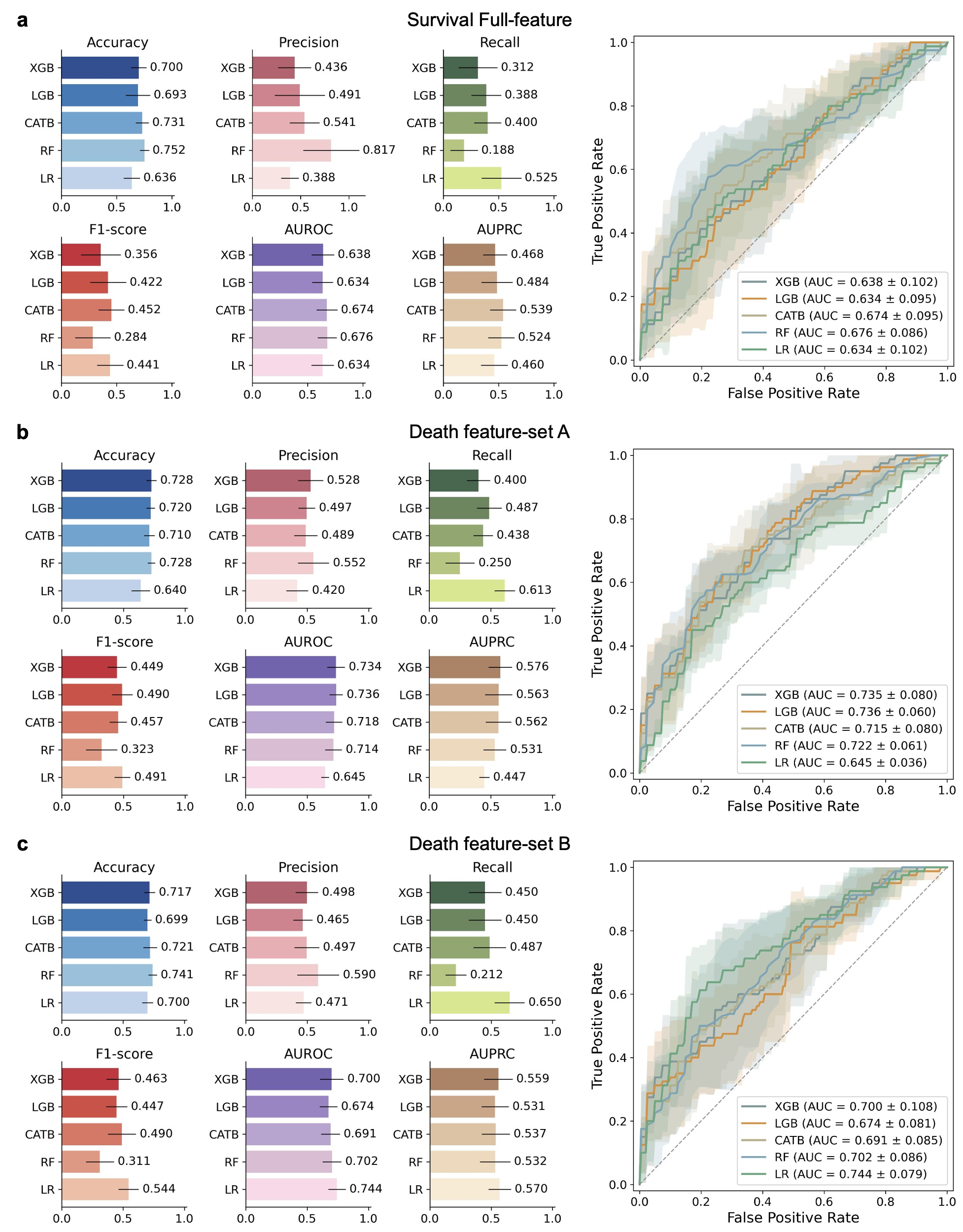

**Supplementary Figure 8. Model performance for 28-day mortality prediction in the CRPA cohort in the MIMIC-IV dataset.**

(a) Overall performance of FFMs for clinical efficacy prediction.

(b-c) ROC curves and performance metrics of the two feature subsets across SFMs.

**Supplementary Table 1.** Baseline characteristics of the study population, including demographics, overall clinical status and comorbidity burden.

|  | Training Cohort  (n = 441) | Temporal Cohort  (n = 111) | p-value |
| --- | --- | --- | --- |
| **Demographics** | | | |
| Age (year) | 66.67 ± 15.97 | 64.71 ± 16.09 | 0.1718 |
| Gender (male, %) | 310 (70.3%) | 86 (77.5%) | 0.1663 |
| Height (cm) | 166.96 ± 7.96 | 167.34 ± 7.97 | 0.3632 |
| Weight (kg) | 63.48 ± 12.73 | 62.77 ± 13.45 | 0.7206 |
| **Setting** | | | |
| Intensive Care Unit (n, %) | 383 (86.8%) | 96 (86.5%) | 1 |
| **Comorbidities** | | | |
| Diabetes Mellitus (n, %) | 146 (33.1%) | 39 (35.1%) | 0.7701 |
| Hypertension (n, %) | 236 (53.5%) | 53 (47.7%) | 0.3266 |
| Heart Disease (n, %) | 216 (49.0%) | 51 (45.9%) | 0.6416 |
| Stroke (n, %) | 150 (34.0%) | 33 (29.7%) | 0.4568 |
| Malignant Tumor (n, %) | 70 (15.9%) | 21 (18.9%) | 0.5287 |
| Chronic Kidney Disease (n, %) | 101 (22.9%) | 17 (15.3%) | 0.1067 |
| Chronic Liver Disease (n, %) | 66 (15.0%) | 21 (18.9%) | 0.3811 |
| COPD (n, %) | 31 (7.0%) | 11 (9.9%) | 0.4106 |
| Other (n, %) | 3 (0.7%) | 3 (2.7%) | 0.0991 |
| **Immunosuppression** | | | |
| Use immunosuppressive agents (n, %) | 64 (14.5%) | 15 (13.5%) | 0.9069 |
| Neutrophil Reduction (n, %) | 6 (1.4%) | 0 (0.0%) | 0.6055 |
| HIV/AIDS (n, %) | 0 (0.0%) | 0 (0.0%) | 1 |
| Post-Transplant Status (n, %) | 11 (2.5%) | 1 (0.9%) | 0.4751 |
| Chemotherapy/Radiation (n, %) | 58 (13.2%) | 18 (16.2%) | 0.4944 |
| Other (n, %) | 310 (70.3%) | 77 (69.4%) | 0.9407 |
| **Infections** | | | |
| HAP (n, %) | 159 (36.1%) | 34 (30.6%) | 0.3372 |
| VAP (n, %) | 282 (63.9%) | 77 (69.4%) | 0.3372 |

**Supplementary Table 2.** Optimized feature subsets identified in the training cohort for each prediction task. For each task, the table lists the representative feature subsets retained after the feature-set selection procedure, the evaluation metric(s) used to define each subset, the number of included variables, and the specific variables retained in the final subset. Abbreviations: ALB, albumin; ALC, absolute lymphocyte count; ALT, alanine aminotransferase; AST, aspartate aminotransferase; CRP, C-reactive protein; eGFR, estimated glomerular filtration rate; FiO₂, fraction of inspired oxygen; HAP, hospital-acquired pneumonia; NEUT%, neutrophil percentage; PCT, procalcitonin; PLT, platelet count; TBIL, total bilirubin； VAP, ventilator-associated pneumonia; WBC, white blood cell count.

| **Task** | **Feature-set ID** | **Optimized metric(s)** | **Included variables** |
| --- | --- | --- | --- |
| Survival | feature-set A | Precision  AUROC | PLT, eGFR, Comorbidities, PCT, Age, ALB, NEUT%, FiO₂, Baseline creatinine |
| Survival | feature-set B | Accuracy  Recall  F1-score  AUPRC | Comorbidities, PLT, Age, NEUT%, CRP, FiO₂, Immunosuppression, Baseline creatinine |
| Clinical efficacy | feature-set A | Accuracy  Recall  F1-Score | FiO_2_, Comorbidities, eGFR, PLT, CRP, Baseline creatinine, Age, TBIL |
| Clinical efficacy | feature-set B | AUROC | FiO_2_, Fungal coinfection, Cefoperazone-sulbactam resistance, HAP, VAP, Kanamycin resistance, eGFR |
| Clinical efficacy | feature-set C | Precision | FiO_2_, Cefoperazone-sulbactam resistance, HAP, VAP, eGFR |
| Clinical efficacy | feature-set D | AUPRC | FiO_2_, Cefoperazone-sulbactam resistance, HAP, Kanamycin resistance, eGFR |
| Polymyxin resistance | feature-set A | AUPRC | Comorbidities, Age, PLT, AST, ALB, CRP, NEUT% |
| Polymyxin resistance | feature-set B | AUROC | Comorbidities, Age, PLT, ALB, WBC, CRP, NEUT% |
| Polymyxin resistance | feature-set C | Accuracy | Comorbidities, Age, AST, PLT, CRP, WBC, NEUT% |
| Polymyxin resistance | feature-set D | Precision  F1-score | Comorbidities, Age, WBC, ALC |
| Polymyxin resistance | feature-set E | Recall | PLT, CRP |
| Treatment duration | feature-set A | MSE | PLT, ALC, Respiratory support level, BMI, ALT |

**Supplementary Table 3.** Optimized feature subsets identified in the CRPA cohort in the MIMIC-IV dataset for each prediction task. For each task, the table lists the representative feature subsets retained after the feature-set selection procedure, the evaluation metric(s) used to define each subset, the number of included variables, and the specific variables retained in the final subset.

| **Task** | **Feature-set ID** | **Optimized metric(s)** | **Included variables** |
| --- | --- | --- | --- |
| Survival | feature-set A | Precision  F1-score  AUROC | pt_worst_4d, lactate_worst_4d, rr_worst_4d, creatinine_worst_4d, age, rdw_worst_4d, bicarbonate_worst_4d |
| Survival | feature-set B | Accuracy  Recall  AUPRC | bicarbonate_worst_4d, age, aptt_worst_4d, ast_worst_4d, respiratory support, creatinine_worst_4d, hr_worst_4d |

**Supplementary Video 1. End-to-end workflow demonstration of Dr.BUG.**

The video illustrates the main workflow of Dr.BUG, including dataset upload, model development, feature-set selection, model release, task tracking, model registry, single-patient prediction, SHAP-based patient-level explanation, batch prediction, regimen-library construction and individualized antibiotic regimen recommendation.
